# Longitudinal Receptive-Expressive Language Profiles in Young Autistic Children

**DOI:** 10.64898/2026.06.02.26354680

**Authors:** Kenza Latrèche, Michel Godel, Fiona Journal, Nada Kojovic, Marie Schaer

**Affiliations:** Autism Brain and Behavior Lab, Faculty of Medicine, University of Geneva, Geneva, Switzerland; Division of Adult Psychiatry, Department of Psychiatry, University Hospitals of Geneva, Switzerland; Department of Psychiatry, University School of Medicine, Geneva, Switzerland; Fondation Pôle Autisme, Geneva, Switzerland

**Keywords:** autism, early childhood, longitudinal design, expressive language, receptive language, language profile, language gap, discrepant profiles

## Abstract

**Background & Aims:** Language development in autism is heterogeneous and strongly predicts later functioning. The balance between receptive and expressive abilities and their developmental trajectories, however, remains poorly understood. While some autistic children exhibit a relative expressive advantage (ExpAdv), others show receptive advantage (RecAdv) or a balanced profile. Prior studies report inconsistent findings and are often limited by cross-sectional designs and small samples. The present study aimed to (1) describe longitudinal trajectories of receptive and expressive language in autistic and typically developing (TD) children; (2) classify children into ExpAdv, Balanced, and RecAdv profiles across early childhood; and (3) examine the stability and transitions of these profiles over time, including associated clinical features.

**Methods:** We analyzed 1,174 longitudinal time points from 318 autistic children and 294 time points from 108 TD children (1.2–5.8 years) from the Geneva Autism Cohort. Receptive and expressive language were assessed with the Mullen Scales of Early Learning. Receptive-expressive balance was quantified as the ratio of receptive to expressive age equivalent scores, classifying children into ExpAdv, Balanced, and RecAdv profiles using adapted cut-offs. Mixed-effects models examined developmental trajectories, and Sankey diagrams visualized profile transitions. Autism features and adaptive behavior were compared across profiles.

**Results:** Autistic children displayed lower expressive and receptive language than TD peers, with receptive abilities exceeding expressive skills in both groups. Overall, 30–35% of autistic children were classified as ExpAdv at 18–36 months, declining to ∼12% by 48–54 months, while Balanced and RecAdv profiles became more prevalent with age. ExpAdv was associated with slower verbal and non-verbal developmental gains. Balanced and RecAdv profiles showed moderate stability (50–60%), whereas ExpAdv was less stable and frequently transitioned toward Balanced profiles. Autistic children with stable ExpAdv profiles were more often female and showed weaker adaptive communication.

**Conclusions:** Receptive-expressive language profiles in autistic children are dynamic. ExpAdv profile is more frequent in younger autistic children, less stable, and linked to slower verbal and non-verbal development and greater autism severity.

**Implications:** ExpAdv may represent an early marker of autism associated with slower expressive and receptive language growth. Longitudinal monitoring of receptive and expressive skills is essential, as transitions toward Balanced or RecAdv profiles are associated with improved developmental outcomes. We discuss clinical considerations regarding language assessment, longitudinal monitoring, and interpretation of receptive-expressive profiles.

## Introduction

Early language development is a strong predictor of long-term outcomes in autistic individuals, including quality of life and independence (Friedman et al., 2019; Howlin et al., 2014; Kover et al., 2016; Loucas et al., 2008). Language acquisition in autism is highly heterogeneous, with substantial variability in level, rate of acquisition, and developmental trajectory (Gernsbacher et al., 2016; Pickles et al., 2014). As language abilities tend to stabilize after age six, the preschool years represent a critical window for identifying early markers of language development and for implementing interventions aimed at supporting language acquisition (Dawson, 2008; Lombardo et al., 2021; Pickles et al., 2014).

Beyond heterogeneous language trajectories in autism, variability in language development also extends to the balance between receptive and expressive skills (Cohenour et al., 2026; Kinard et al., 2026; Reinhartsen et al., 2019). In typical development, receptive language reliably precedes expressive language and develops much more rapidly (Benedict, 1979; Gernsbacher et al., 2016; Gershkoff-Stowe & Hahn, 2013; McDaniel et al., 2018; Ryan et al., 2016), with children understanding words before they are able to produce them. In autism, however, the dynamic of receptive-expressive development may vary substantially. Some children show relatively stronger expressive than receptive skills (Expressive Advantage; ExpAdv), others the reverse (Receptive Advantage; RecAdv), and others a broadly balanced profile (Cohenour et al., 2026; Davidson & Ellis Weismer, 2017; Kinard et al., 2026; Reinhartsen et al., 2019). Importantly, these three profiles are independent of overall language ability and do not indicate language delay; children with significant language impairments may nevertheless exhibit a balanced receptive-expressive profile (Cohenour et al., 2026). Consequently, children with comparable overall language levels may show markedly different receptive and expressive skill patterns, potentially leading to distinct developmental trajectories (Cohenour et al., 2026). Indeed, children with a relative ExpAdv at baseline showed slower expressive gains over one year compared to those with a RecAdv or Balanced profile (Cohenour et al., 2026). This suggests that receptive-expressive balance may may provide information about early language development that can be obscured when receptive and expressive abilities are considered only as an aggregated measure.

However, the literature on receptive-expressive profiles in autism remains inconsistent (McDaniel et al., 2018). Prevalence estimates for ExpAdv range from 24% to 43%, across autistic samples, compared to approximately 11% in typically developing (TD) children (Cohenour et al., 2026; Reinhartsen et al., 2019; Seol et al., 2014). Evidence on age-related effects is also mixed: some studies reported a stronger expressive advantage in younger children that diminishes with age (Davidson & Ellis Weismer, 2017; Hudry et al., 2010; Kinard et al., 2026; Reinhartsen et al., 2019; Seol et al., 2014) whereas others found no significant age effects (Cohenour et al., 2026). A meta-analysis of 74 studies reported comparable impairments in receptive and expressive language overall, although most studies directly comparing both domains did identify a receptive-expressive gap (Kwok et al., 2015). These inconsistencies likely reflect differences in how profiles are operationalized. Some studies divided receptive by expressive age equivalent scores and used either an arbitrary cutoff or one based on TD samples to define the three profiles (ExpAdv, RecAdv, Balanced; Cohenour et al., 2026; Kinard et al., 2026; Reinhartsen et al., 2019; Seol et al., 2014), while others computed the difference between receptive and expressive standard scores, with negative values indicating an ExpAdv profile and positive values indicating a RecAdv profile (Davidson & Ellis Weismer, 2017). Because these approaches rely on different metrics, scales, and classification thresholds, they may assign children differently across receptive-expressive profiles, leading to differences in estimated prevalence and limiting direct comparisons across studies. Other studies compared receptive and expressive vocabulary counts without defining distinct profile categories (Hudry et al., 2010; McDaniel et al., 2018). In addition to these methodological differences, inconsistent findings may reflect instability in receptive-expressive profiles across development (McDaniel et al., 2018), which cannot be fully captured by cross-sectional designs.

Longitudinal studies are essential for characterizing the developmental stability and transitions of receptive-expressive language profiles, yet such evidence remains scarce. Although existing studies suggest that receptive-expressive gaps may decrease over time and that ExpAdv may predict slower expressive growth (Cohenour et al., 2026; Davidson & Ellis Weismer, 2017), profiles are typically assessed at a single time point, leaving their stability and dynamics poorly understood. In previous work, we identified three distinct language trajectory subgroups in autistic children (Language Unimpaired, Language Impaired, and Minimally Verbal), demonstrating that autistic children differ not only in language level but in their developmental course (Latrèche et al., 2024). The present study extends that investigation by examining whether receptive-expressive balance provides additional predictive value beyond overall language proficiency.

Beyond characterizing these developmental trajectories, it is also important to consider whether early intervention influences receptive-expressive language balance specifically, rather than overall language level alone. Early intensive intervention programs, such as the Early Start Denver Model (ESDM), have been shown to improve language and cognitive outcomes in young autistic children (Dawson et al., 2010; Rogers et al., 2019; Wang et al., 2022). However, it remains unknown whether ESDM also affects the balance between receptive and expressive language, leading to shifts across ExpAdv, RecAdv, or Balanced profiles. Given that ExpAdv may reflect a developmentally transient but clinically informative imbalance, understanding whether early intervention relates to the resolution of this imbalance could have direct relevance for how intervention outcomes are evaluated.

Leveraging a longitudinal dataset comprising 1,174 time points from 318 autistic children (1.2–5.8 years) and 294 time points from 108 TD children (1.2–5.7 years), we aimed to (1) characterize group-level receptive and expressive language trajectories; (2) identify ExpAdv, RecAdv, and Balanced profiles across early childhood by classifying children at multiple age windows; and (3) assess profile stability, transitions, and predictors, such as enrollment in early intervention (ESDM). We hypothesized that ExpAdv would be more prevalent in younger autistic children and associated with slower language growth, positioning the receptive-expressive balance as a clinically meaningful early marker with implications for prognosis and intervention.

## Methods

### Participants

The present study drew participants from the ongoing Geneva Autism Cohort, in which autistic children and typically developing (TD) peers are assessed every six months over two years. For autistic participants, diagnosis was confirmed by a child psychiatrist (MS) according to DSM-5 criteria (American Psychiatric Association, 2013). All autistic participants underwent the Autism Diagnostic Observation Schedule (ADOS-G/ADOS-2; (Lord et al., 2000, 2012) and the Autism Diagnostic Interview-Revised (ADI-R; Lord et al., 1994), a semi-structured parent interview assessing autism features, and scored above diagnostic cut-offs. Before inclusion in the cohort, TD children also underwent the ADOS-G/ADOS-2 and the ADI-R to confirm the absence of autistic features. TD children were additionally screened for any developmental concern, somatic and psychiatric concerns, and for the absence of autism in first-degree family members. Development of TD and autistic children was assessed using the Mullen Scales of Early Learning (MSEL; Mullen, 1995). For more information about the Geneva Autism Cohort, see (Latrèche et al., 2024). From the longitudinal cohort, we included all participants who had at least one available MSEL assessment. Our final sample comprised 426 participants and 1,468 time points. The autism spectrum disorder (ASD) group was composed of 318 children (62 females, age range: 1.2 to 5.8 years, 1,174 time points), and the TD group included 108 children (42 females, age range: 1.2 to 5.7 years, 294 time points). Sample characteristics are presented in Table 1.

**Table 1.**
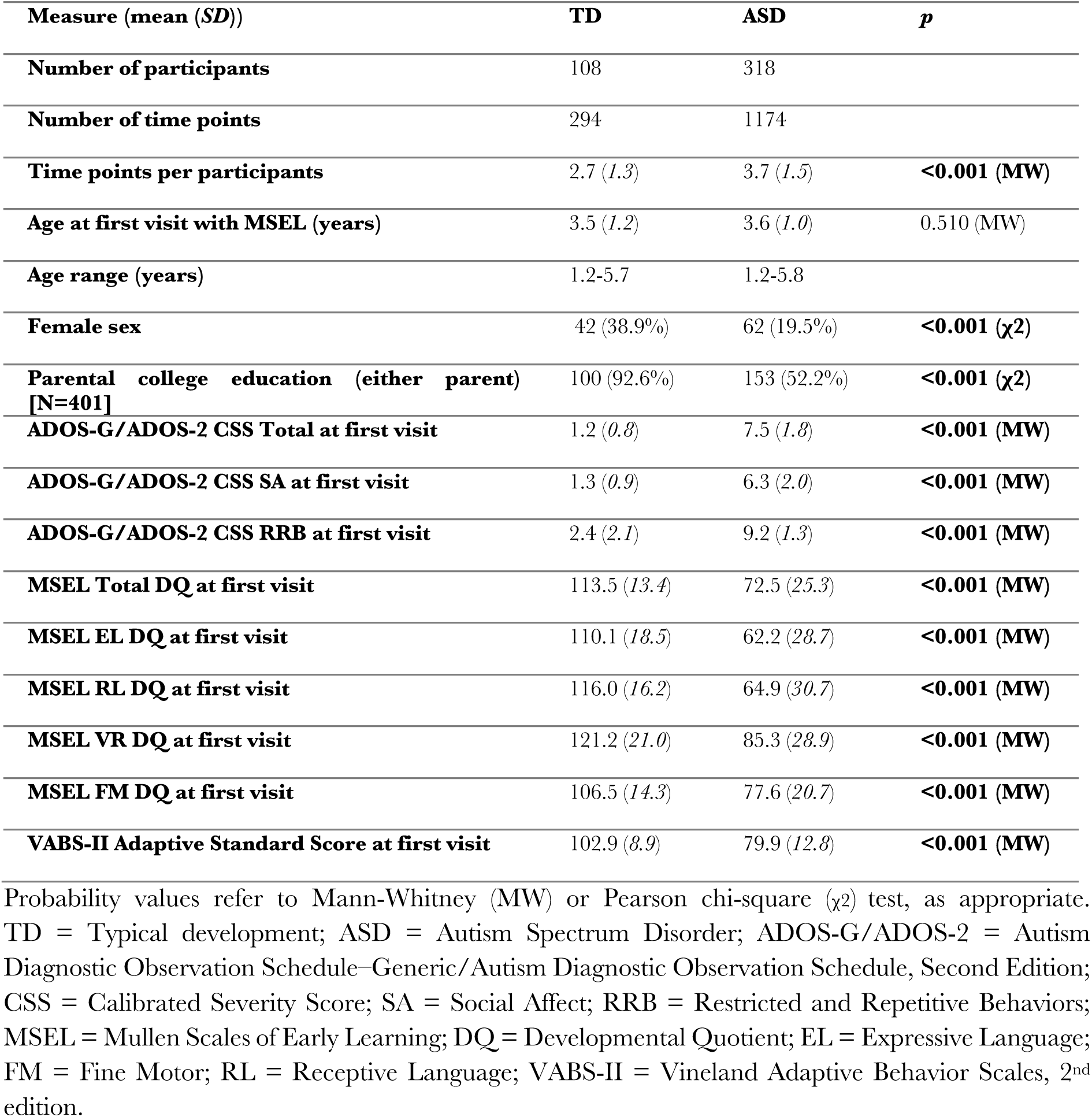
Sample characteristics of the TD and ASD groups.

### Measures

We collected several types of measures. First, verbal and non-verbal developmental quotients (DQs) were used both to characterize the sample and as primary outcome measures in longitudinal analyses. Second, receptive language and expressive language measures were used to define the three language profiles (ExpAdv, Balanced, RecAdv), which served as the main grouping variable for comparisons between the autistic and TD groups, as well as for intra-group longitudinal analyses. Third, autism features and adaptive skills were used as descriptive clinical measures, and as markers to examine profile stability and transitions over time. Finally, we examined whether early intervention was associated with a higher probability of transitioning between profiles over time.

### Development

The MSEL assesses development in four domains: Fine Motor (FM), Visual Reception (VR), Receptive Language (RL) and Expressive Language (EL). Developmental quotients (DQs) were computed for each domain and for composite non-verbal (FM and VR) and verbal (RL and EL) domains by dividing age equivalent scores by the child’s chronological age and multiplying by 100 (Lord et al., 2006). Because floor effects are frequent when using MSEL T-scores in children with significant developmental delays (Kinard et al., 2026), DQs may offer a more informative estimate of developmental level by preserving inter-individual variability among lower-performing participants.

To measure the receptive-expressive language balance, we computed ratios by dividing RL by EL age equivalent scores. Previous studies that used this ratio (Cohenour et al., 2026; Seol et al., 2014) have categorized the three profiles using canonical cut-offs: (1) Receptive Advantage: ratio above 1.10; (2) Balanced: ratio between 0.90 and 1.10; (3) Expressive Advantage: ratio below 0.90. Another study (Reinhartsen et al., 2019) observed that their TD group (N = 889, 2.5–5.7 years) exhibited higher RL skills than EL skills when assessed with the MSEL. Thus, the authors used the mean (1.04) and standard deviation (0.13) of their TD sample to adapt the ratio cut-offs: (1) Receptive Advantage: ratio above 1.17; (2) Balanced: ratio between 0.91 and 1.17; (3) Expressive Advantage: ratio below 0.91. This adapted ratio accounts for the MSEL tendency to yield higher RL scores compared to EL scores. In our TD longitudinal sample, the mean ratio was 1.13, and the standard deviation was 0.21, showing an even greater skewness toward RL. Since the sample of Reinhartsen and colleagues was more robust than our sample in terms of sample size (N = 889), with a similar age range (2.5-5.7 *vs.* 1.2–5.7 years), we used their adapted cut-offs (Reinhartsen et al., 2019).

### Autism features

All participants underwent the Autism Diagnostic Observation Schedule-Generic (ADOS-G; (Lord et al., 2000) or the second edition (ADOS-2; (Lord et al., 2012). We administered the modules of the ADOS-G/ADOS-2 based on the individual’s age and level of language (Toddler Module, and Modules 1, 2, and 3). Calibrated Severity Scores (CSS) for Total score, Social Affect, and Restricted and Repetitive Behaviors were used to compare symptom severity across modules (Gotham et al., 2009).

Item-level scores were also extracted across modules to examine autistic features across profiles in a supplementary analysis.

### Adaptive skills

The Vineland Adaptive Behavior Scales, second edition (VABS-II; Sparrow et al., 2005) were administered to all parents longitudinally to measure adaptive skills in Communication, Daily Living skills, Socialization, and Motor skills. Standard scores (mean = 100, SD = 15) were used.

### Early Intervention

Within the autistic group, 188 children (763 time points) were enrolled in a 2-year early intensive intervention program based on the Early Start Denver Model (ESDM; Rogers, 2016). The ESDM belongs to Naturalistic Developmental Behavioral Interventions, which are evidence-based approaches combining principles of operant learning and developmental science to encourage acquisition of new skills in naturalistic contexts (Schreibman et al., 2015; Vivanti & Stahmer, 2021). The intervention is individualized (one therapist per child) and the intensity varies between 15–20 hours per week (for more details, see Godel et al., 2022).

### Statistical Analyses

#### Sliding-window classification of language profiles

As our longitudinal dataset spans from toddlerhood to childhood, the autistic group was divided into age windows using a sliding-window approach ranging from 18 to 54 months (6-month width, 1-month increment; e.g., 18–24, 19–25 months). Age windows were defined with the lower bound included and the upper bound excluded (e.g., a child aged exactly 24 months was included in the 24–30-month window, not the 18–24-month window). Within each window, children were assigned to a profile (ExpAdv, Balanced, RecAdv) based on the ratio (Reinhartsen et al., 2019). If a child had multiple time points within the same window, the earliest was selected, though this was rare given that our protocol aims to target six-month assessment intervals (ASD: 0–8.6% of subjects per age window had >1 time point, M = 3.7%; TD: 0–7.5%, M = 1.8%). All available time points for each child, prior and subsequent to the classification, were included in mixed-effects modeling. Due to the number of statistical tests, these profile-distribution comparisons were interpreted descriptively.

To compare profile distributions between the autistic and TD longitudinal datasets, analyses were restricted to visits between 18 and 54 months of age (consistent with intra-group analyses), yielding 783 visits from autistic children and 203 visits from TD peers. Given that the distribution of visits across age-windows differed between groups, a multinomial logistic regression was used to test the effect of group (ASD *vs*. TD) on profile classification (Balanced, ExpAdv, RecAdv), with age-window included as a covariate. Age-adjusted marginal proportions were estimated using the average marginal effects method.

#### Receptive and expressive language trajectories

First, to describe receptive and expressive language trajectories in the autistic and TD groups, we conducted mixed-effects analyses. Mixed-effects modeling is a well-suited approach for longitudinal datasets with varying numbers of time points per participant (Latrèche et al., 2024; Mutlu et al., 2013). Age and group (ASD, TD, ExpAdv, Balanced, RecAdv) were included as fixed effects along with their interaction term. Participant-specific random slopes for age accounted for individual variability in developmental change. Analyses were conducted using the myMixedModelsTrajectories toolbox (available at https://github.com/danizoeller/myMixed-ModelsTrajectories) in MATLAB® R2019b (MathWorks, Natick, MA). Linear and quadratic random-slope models were fit to capture different age-related developmental patterns. The best fitting model was selected according to the Bayesian information criterion. Behavioral outcomes included EL and RL DQs and age equivalent scores. Model fit was evaluated using the Bayesian Information Criterion, yielding, e.g., a quadratic model as follows:

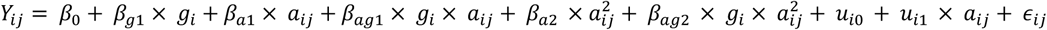

*Y* : *RL Developmental Quotient or Age Equivalent Score, EL Developmental Quotient or Age Equivalent Score, Verbal Developmental Quotient, Non-verbal Developmental Quotient*

*i,j: [subject, timepoint] index*

*β_xn_* : *fixed effects*

*g* : *grouping variable*

*α* : *age*

*u* : *normally distributed random effect*

*∈_ij_* : *normally distributed error term*

Second, to determine developmental outcomes in each of the autistic language profiles (ExpAdv, Balanced, and RecAdv), we applied the same modeling framework across the three autistic language profiles (ExpAdv, Balanced, and RecAdv) across sequential age windows defined in the above section. Behavioral outcomes consisted of MSEL Verbal and Non-Verbal DQs. Given the number of statistical tests involved, these trajectory comparisons were interpreted descriptively, as characterizing overall developmental patterns across profiles, rather than as a strictly confirmatory set of tests.

#### Stability and transitions of language profiles

To examine each language profile’s stability, we focused on the six main age windows (i.e., 18–24, 24– 30, 30–36, 36–42, 42–48, 48–54 months). Transitions between profiles were visualized using Sankey diagrams. For each window-to-window transition, autistic children who remained in the ExpAdv profile were labeled ‘ExpAdv-Stable’, while those who shifted to Balanced and RecAdv profiles were labeled ‘ExpAdv-Switch’. These subgroups were compared on sex distribution, enrollment in early intervention, and adaptive skills. For the post hoc comparison of ExpAdv-Stable and ExpAdv-Switch subgroups at each transition, a Bonferroni correction was applied across the six baseline variables tested within each transition (sex distribution, early intervention, and the four VABS adaptive domains; α = 0.05/6 = 0.0083).

#### Supplementary ADOS-G/ADOS-2 analyses

To further describe differences in autism features across language profiles, autistic children were classified into ExpAdv, Balanced, and RecAdv at their earliest time point, corresponding to diagnostic confirmation assessments. In this cross-sectional sample, profiles were compared on ADOS-G/ADOS-2 calibrated severity scores (Total, Social Affect, and RRB; Gotham et al., 2009) and at the item level.

## Results

### Description of TD and autistic groups

The 108 typically developing (TD) participants (294 time points) and the 318 autistic participants (1,174 time points) did not differ in age at the first time point (mean ± SD: TD = 3.5 ± 1.2; ASD = 3.6 ± 1.0 years) (Table 1). The TD group (38.9%) included more females than the autistic group (19.5%) and parents of TD children had higher education than parents in the autistic group. At baseline, autistic participants showed greater autism symptom severity, lower non-verbal and verbal skills, and reduced adaptive functioning compared with TD peers.

The baseline differences of sex distribution and parental education motivated two complementary analytic strategies. For within-group analyses (developmental trajectories within each group, profile stability and transitions, and predictors of transition), we retained the full, representative ASD (N=318) and TD (N=108) samples to preserve statistical power, since these analyses do not involve a direct ASD-TD comparison. For the two analyses that do directly compare ASD and TD (developmental trajectory group differences and profile-distribution differences), we additionally verified our findings in a subsample matched on sex and parental education, drawn independently 10 times to ensure stability (see Table S1). Findings from the matched analyses were consistent with the full-sample results.

### Trajectories of receptive and expressive language

Expressive and receptive language age equivalent scores were significantly lower in the autistic group than in the TD group (*p* <0.001), with a significant age × group interaction indicating distinct developmental trajectories (*p* <0.001) (Table S2, Figure 1A–B). This indicates both lower language levels and slower gains in autistic children. Within the autistic group, receptive skills were higher than expressive language skills (*p* <0.001) (Figure 1C), a pattern also seen in TD children (*p* <0.001) (Figure 1D). Comparable group differences and developmental trajectories were observed for DQs (Figure S1).

**Figure 1.**
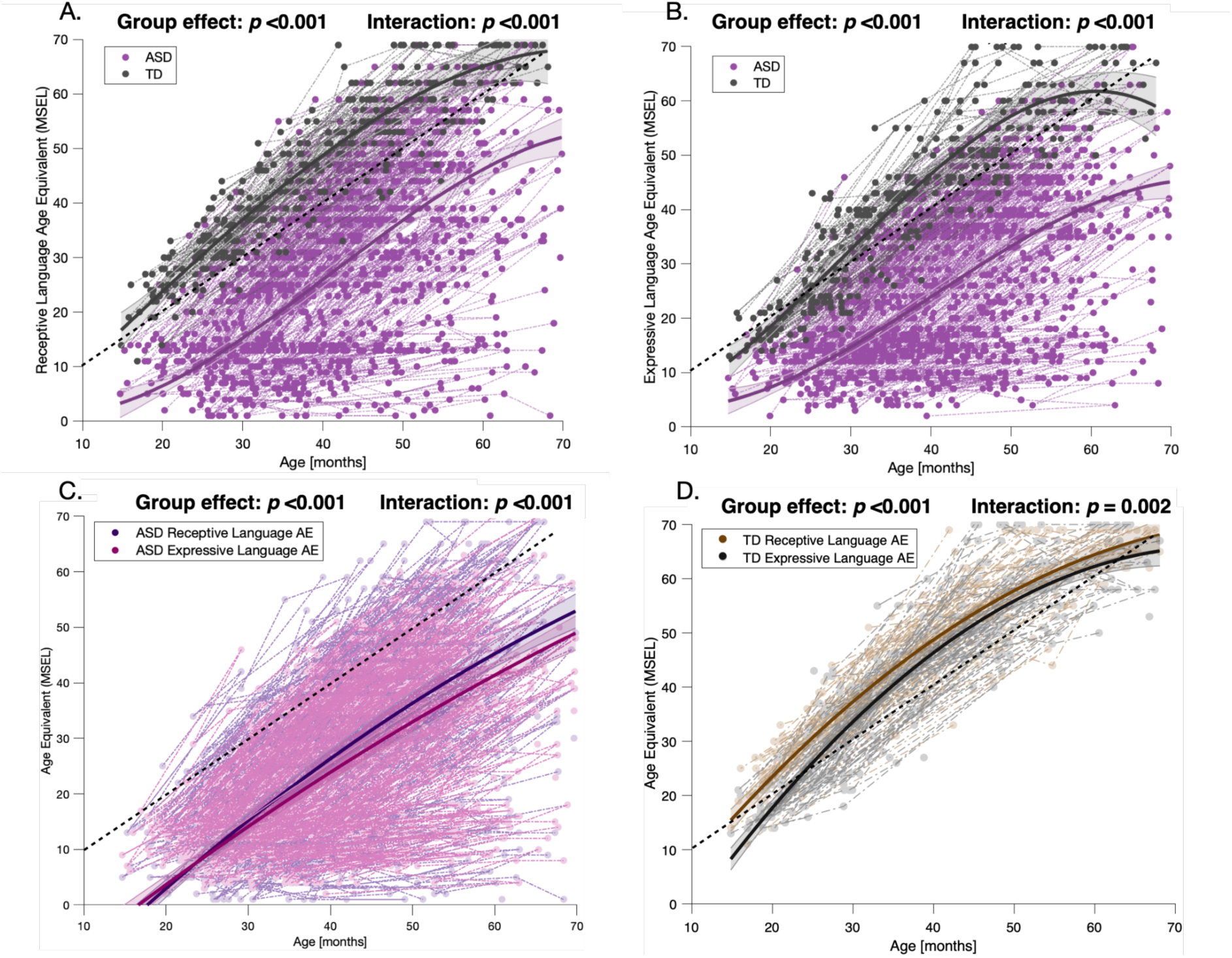
Developmental trajectories of receptive and expressive language age equivalents (MSEL) across age in the ASD and TD groups. (A) Receptive Language trajectories in ASD and TD participants. (B) Expressive Language trajectories in ASD and TD participants. (C) Overlay of receptive and expressive language trajectories within the ASD group. (D) Overlay of receptive and expressive language trajectories within the TD group. The colored bands around the estimated group-level trajectory indicate the 95% confidence interval. MSEL : Mullen Scales of Early Learning; ASD : Autism Spectrum Disorder; TD: typical development; AE : Age Equivalent Score.

Results on the 10 sample draws matched on sex and parental education were highly consistent with those obtained in the full sample: autistic children showed lower expressive and receptive language development compared with TD children (main group effect significant in 10/10 draws); and the age × group interaction was significant in 10/10 draws for expressive language, and 7/10 draws for receptive language (Table S3).

### Sliding-window classification of language profiles

The distribution of language profiles differed significantly between the autistic and TD groups after controlling for age-window (LR χ²(2) = 30.87, *p* <0.001, see Figure S2 and Table S4). The ExpAdv profile was more common in autism (26%) than TD (10%; OR = 3.61 [2.13–6.13], *p* <0.001), and Balanced profiles were more frequent in TD (48%) than autism (41%; OR = 0.66 [0.48–0.92], *p* = 0.013). RecAdv proportions did not differ significantly between groups after age adjustment (ASD: 33%, TD: 42%; OR=1.04 [0.73–1.47], *p* = 0.844). Thus, autistic children had 3.6 times the odds of presenting an ExpAdv profile compared with TD peers, whereas TD children had 1.5 times the odds of presenting a Balanced profile than autistic children. RecAdv was equally distributed across groups once age was considered.

This pattern was similarly preserved across the 10 matched-sample draws (Table S5): autistic children showed significantly higher odds of an ExpAdv profile than TD peers in 9 of 10 draws (OR range: 1.56– 2.48), and RecAdv proportions did not differ in any draw. The relative increase in Balanced profiles among TD children was directionally consistent across all 10 draws but reached significance in only 5, likely reflecting reduced power in a subsample retaining roughly a quarter of the original autistic cohort.

The autistic group (N = 318, 1,174 time points) was then divided into 6-month windows from 18 to 54 months, and children were assigned profiles based on the ratio (Figure 2, Table S6). ExpAdv prevalence was ∼30–35% between 18–36 months, declining to ∼12% by 48–54 months. Conversely, the Balanced profile increased from ∼35% to ∼50%, while RecAdv rose modestly to ∼35% by 48–54 months. These results indicate that autistic children increasingly display a Balanced profile, with fewer showing a pronounced expressive advantage in later toddlerhood. Sliding-window results for TD children are shown in Figure S3 (Table S7): RecAdv decreased from ∼80% to ∼20% between 18 months and 54 months, while Balanced increased and ExpAdv remained rare.

**Figure 2.**
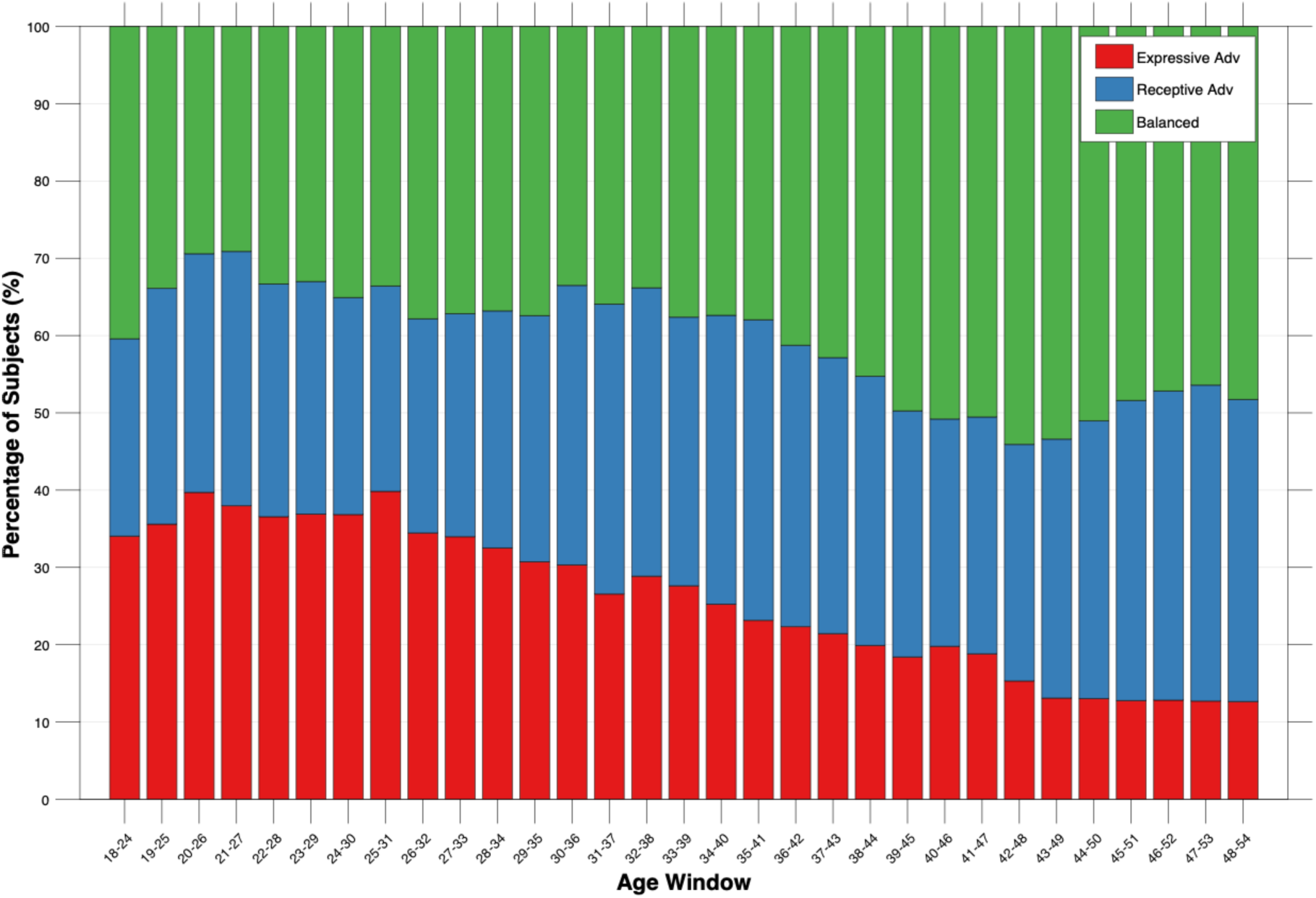
Proportion of Expressive Advantage, Balanced, and Receptive Advantage profiles within each age window in the ASD group.

Within the autistic group, the three profiles differed significantly in their developmental trajectories for expressive language (Figure S4), receptive language (Figure S5), the aggregate verbal measure (Figure S6), and the non-verbal domain (Figure S7) across the six age windows (group effect *p*s = <0.001–0.006). Group × age interaction effects (*p*s = <0.001–0.535) were more consistently significant at older age windows. ExpAdv children exhibited consistently lower verbal and non-verbal trajectories compared to RecAdv and Balanced profiles, which displayed similar developmental levels.

### Stability and transitions of language profiles

To examine profile stability, we analyzed consecutive transitions across six age windows, computing the percentage of children who remained in the same profile or who switched to another profile. Stability and transition patterns are shown in Figure 3 and Table S8.

**Figure 3.**
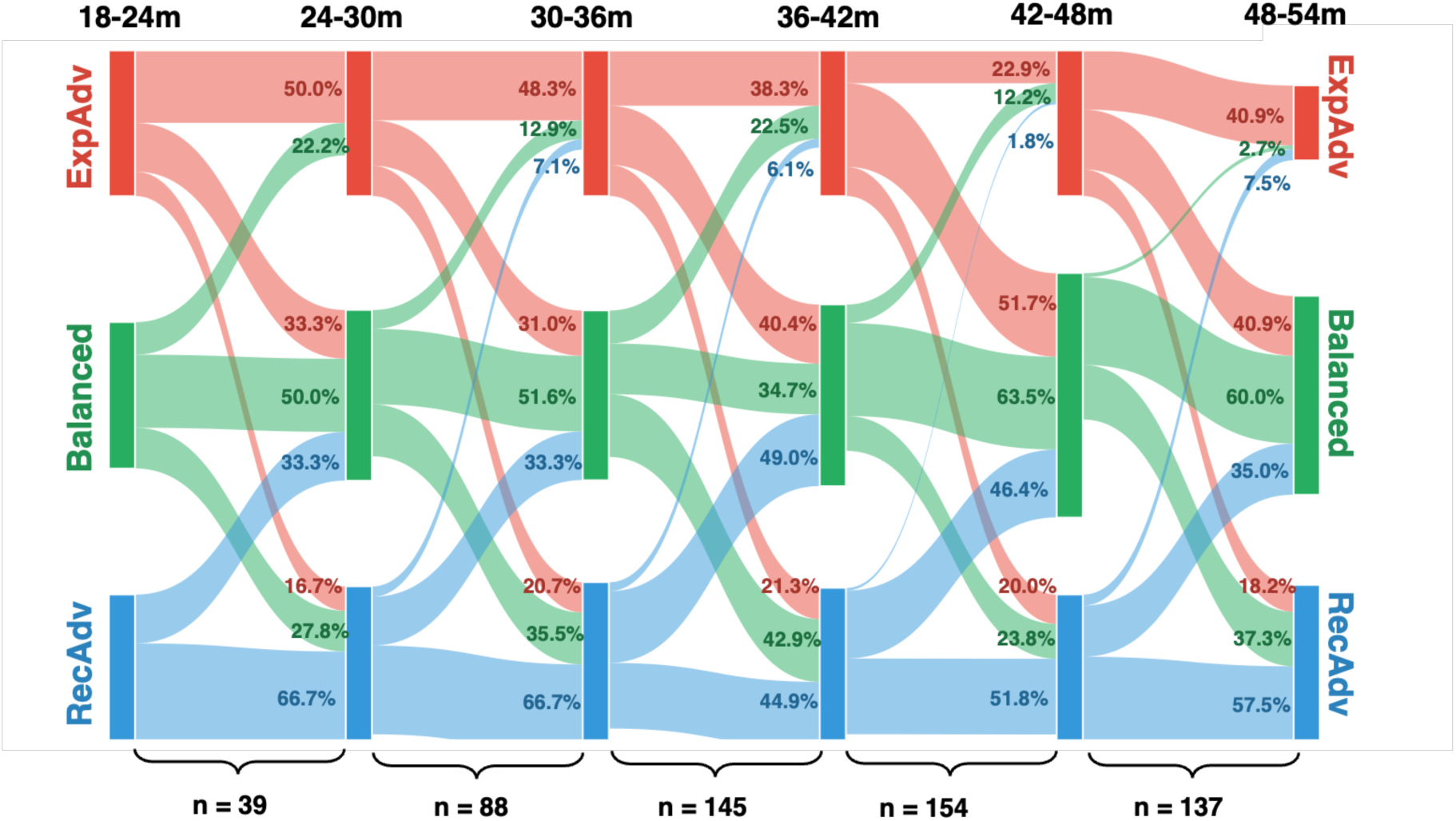
Sankey diagram illustrating stability and transitions of Expressive Advantage (red), Balanced (green), and Receptive Advantage (blue) profiles across six age windows (18–54 months) in the autistic group. Each vertical bar represents the proportion of children in each profile at a given age window, and the widths of each flow reflect the number of children who remained in the same profile or transitioned to another. Percentages indicate the proportion of children moving from each source profile to each destination profile. Sample sizes below each transition reflect the number of children with data available at both consecutive time points.

Balanced and RecAdv profiles showed moderate stability over development, with ∼50–60% of children remaining in the same profile across consecutive age windows (Figure 3). Transitions between the Balanced and RecAdv were relatively common (∼30–50% of participants), suggesting that these two profiles reflect closely related developmental patterns. In contrast, transitions from either Balanced or RecAdv into ExpAdv were uncommon (∼0–20%). Overall, the ExpAdv profile showed markedly lower stability than the other profiles. Across development, many children initially classified as ExpAdv transitioned toward Balanced (∼30–50%) or, less frequently, RecAdv (∼20%) profiles. This instability was particularly apparent before 42 months. After this age, the proportion of children remaining in the ExpAdv profile appeared somewhat more stable (∼40%).

As the ExpAdv profile showed overall limited stability and frequent transitions, we conducted a post hoc analysis to examine predictors of profile transition. For each consecutive window, we compared children with ExpAdv who transitioned to Balanced or RecAdv (ExpAdv-Switch) with those who remained in the ExpAdv profile (ExpAdv-Stable). From 30–36 months to 36–42 months, ExpAdv-Stable (n = 18) had a higher proportion of females and showed weaker adaptive communication than ExpAdv-Switch children (n = 29) (Figure S8), with both differences surviving Bonferroni correction (α = 0.0083). However, these findings should be interpreted cautiously, given that the sex-distribution contrast is driven by a single female in the ExpAdv-Switch group, and both comparisons rely on modest subgroup sizes. A lower proportion of ExpAdv-Stable children had received early intervention (*p* = 0.017), but this association did not survive correction for multiple comparisons and should therefore be interpreted as exploratory. Verbal growth was also slower in ExpAdv-Stable (Figure S9, p <0.001). No significant group differences were observed at the other developmental transitions (Table S9).

### Supplementary analysis: Autism features across language profiles

To further examine clinical features associated with language profiles, we conducted a cross-sectional analysis at each child’s first available time point. ExpAdv children showed higher Total and Social Affect ADOS-G/ADOS-2 severity than RecAdv and Balanced profiles (Figure S10). Moreover, item-level analyses indicated elevated scores in ExpAdv children on language-related items (e.g., stereotyped language, immediate echolalia), specific social-communication skills (e.g., requesting, showing, joint attention), and restricted and sensory interests compared to children with Balanced and RecAdv profiles (Figure S11, Table S10). Overall, children with Balanced and RecAdv profiles demonstrated comparable scores. These results suggest that the ExpAdv profile was associated with greater autism symptom severity at cohort enrollment, particularly in social communication and language domains.

## Discussion

In this study, we investigated receptive-expressive language development in a large longitudinal sample of autistic and typically developing (TD) children. By classifying children into Expressive Advantage (ExpAdv), Receptive Advantage (RecAdv), and Balanced profiles, we characterized developmental trajectories, prevalence, and stability over early childhood. Receptive-expressive language profiles were dynamic and clinically meaningful, with ExpAdv emerging as an early, less stable profile associated with slower verbal and non-verbal cognitive development.

Consistent with prior literature, autistic children exhibited lower receptive and expressive language than TD peers, along with slower developmental gains. Receptive language exceeded expressive language in both groups. While this gap remained stable in TD children, it widened over time in autistic children. This pattern contrasts with previous reports of decreasing receptive-expressive gap with age (Davidson & Ellis Weismer, 2017; Hudry et al., 2010). This discrepancy likely reflects differences in assessment instruments (e.g., parent report versus direct assessment) (Nevill et al., 2019) and in the operationalization of receptive-expressive profiles (e.g., age-equivalent ratios, difference scores, or standard score discrepancies; Cohenour et al., 2026; Davidson & Ellis Weismer, 2017; Hudry et al., 2010; Kinard et al., 2026; McDaniel et al., 2018; Reinhartsen et al., 2019; Seol et al., 2014). The present study relied on the Mullen Scales of Early Learning (MSEL), for which TD children show slightly higher receptive than expressive age equivalent scores on average (Reinhartsen et al., 2019). Consequently, receptive-expressive balance should be interpreted relative to normative MSEL performance. This underscores the importance of adapting ratio cut-offs to both the assessment instrument and normative TD data, rather than relying on canonical thresholds (Cohenour et al., 2026; Seol et al., 2014).

Our findings emphasize that ExpAdv is more characteristic of autism than of typical development. Although not the most prevalent profile overall, it was most common in younger autistic children, in line with previous reports (Kinard et al., 2026; Reinhartsen et al., 2019; Seol et al., 2014). Evidence from studies including children with developmental delay (DD) indicates that ExpAdv is less prevalent in DD than in autism (Kinard et al., 2026; Reinhartsen et al., 2019; Seol et al., 2014). Although these findings suggest that ExpAdv may be more strongly associated with autism, its specificity has yet to be established. Future studies comparing autistic individuals with children with other neurodevelopmental and genetic conditions are needed to determine whether ExpAdv represents an autism-specific phenotype or a broader feature of atypical neurodevelopment.

Importantly, ExpAdv reflects a relative expressive advantage compared to receptive abilities, rather than strong expressive language skills in absolute terms. Indeed, most autistic children within this profile showed global language delay, with receptive difficulties being particularly pronounced. Consistent with this interpretation, ExpAdv was associated with slower receptive and expressive language development as well as reduced non-verbal cognitive growth in autistic children (Cohenour et al., 2026; Reinhartsen et al., 2019). Importantly, this finding indicates that although ExpAdv is characterized by relatively higher expressive skills, the majority of children within this profile show globally lower language abilities, with receptive language representing the more pronounced relative weakness.

Beyond language level, children with an ExpAdv profile at cohort enrollment exhibited greater autism symptom severity, particularly in social communication, and showed RRB severity comparable to the Balanced profile and higher than the RecAdv profile. Item-level analyses further revealed a distinctive social communication profile in children with ExpAdv, characterized by more frequent stereotyped language and immediate echolalia, together with reduced use of gestures and lower rates of social initiation (e.g., requesting, showing, and joint attention). These findings suggest that the relatively higher expressive scores in ExpAdv may partly reflect greater use of stereotyped and echoed forms of language, consistent with previous work (Kinard et al., 2026; Paul et al., 2008). Children with an ExpAdv profile also showed fewer spontaneous vocalizations directed to others across pragmatic contexts, indicating reduced functional communication despite relatively higher expressive language scores. Together, these patterns suggest that children with an ExpAdv profile may demonstrate relatively stronger expressive language skills while continuing to experience difficulties using language effectively in reciprocal social interactions. Extending previous cross-sectional findings, our longitudinal results further show that ExpAdv is not only associated with slower verbal and non-verbal developmental trajectories over time but also with greater social communication difficulties.

Longitudinal analyses also demonstrated that receptive-expressive profiles are dynamic rather than static across development. Although the proportions of RecAdv and Balanced profiles remained moderately stable over time, transitions between these profiles were frequent and bidirectional, suggesting that these profiles reflect a continuum of relatively typical receptive-expressive organization associated with higher language outcomes. Importantly, transitions between profiles in the Sankey diagrams followed structured patterns rather than appearing entirely random, suggesting the presence of meaningful developmental trajectories. Variability between these profiles may be driven by both developmental change and measurement characteristics (consistent with the MSEL yielding relatively higher receptive scores in TD children; Reinhartsen et al., 2019). In contrast, transitions from either RecAdv or Balanced into ExpAdv were uncommon, whereas children classified as ExpAdv frequently transitioned to other profiles, most often to Balanced. Together, these findings suggest that ExpAdv represents a more distinct and developmentally transient profile. Children classified as ExpAdv were more likely to transition out of this profile than children in other profiles were to transition into it. This pattern was characterized by relative delays in both receptive and expressive abilities, with the imbalance appearing to partially resolve over time.

The ExpAdv profile showed declining stability up to 42–48 months, with considerable transitions particularly toward the Balanced profile. From 48–54 months, ExpAdv stability increased, although the limited number of participants older than 54 months prevents conclusions about whether this reflects a meaningful developmental turning point. Future studies including older, school-age participants are needed to determine whether the 42–48-month window constitutes a meaningful developmental shift in receptive-expressive organization. Several mechanisms may contribute to the decline in ExpAdv prevalence with age. First, this pattern may reflect differential developmental trajectories of expressive and receptive language, whereby autistic children with an ExpAdv profile showed slower verbal and non-verbal development overall, with receptive skills appearing disproportionately affected at younger ages. One possibility is that receptive language develops at a relatively faster rate over time, gradually reducing the prevalence of ExpAdv observed early in development in autism. This interpretation is consistent with the general pattern of receptive language preceding expressive language in typical development (Figure 1D; Benedict, 1979; Gernsbacher et al., 2016; Gershkoff-Stowe & Hahn, 2013; McDaniel et al., 2018; Ryan et al., 2016). Thus, although these mechanisms cannot be fully disentangled in the present study, our findings reinforce that receptive-expressive profiles should be interpreted as dynamic developmental patterns rather than fixed characteristics.

Among children who remained in the ExpAdv profile, we observed a higher proportion of girls between 30–36 and 36–42 months, raising the possibility of sex-related differences in receptive-expressive language trajectories. However, as this association was observed at only one developmental transition and involved a small number of girls, it should be interpreted cautiously. Although sex differences in autism presentation have been increasingly recognized, language development across autistic males and females remains largely unexplored (Paolizzi et al., 2026). Whether specific receptive-expressive language profiles, such as ExpAdv, differ by sex remains unclear. Future longitudinal studies with larger, sex-balanced samples of young autistic children are needed to determine whether this preliminary observation reflects a reproducible developmental pattern.

Early intervention may be associated with transitions toward more balanced receptive-expressive language profiles. Between 30–36 and 36–42 months, children who transitioned from ExpAdv to Balanced or RecAdv showed a nominally higher rate of ESDM intervention and faster verbal growth than those who remained in the ExpAdv profile. However, the association with ESDM intervention did not survive correction for multiple comparisons and was observed at only one of five transitions examined. It should therefore be considered exploratory rather than conclusive evidence of an intervention effect, particularly given the non-randomized design of our study. If replicated in independent samples, this finding would provide preliminary support for an association between ESDM exposure and more balanced receptive-expressive profiles, highlighting the potential relevance of considering both overall language level and receptive-expressive balance when evaluating developmental outcomes following early support (Dawson et al., 2010; Latrèche et al., 2024; S. J. Rogers et al., 2019; Wang et al., 2022).

Overall, these findings position receptive-expressive balance as a relevant fine-grained dimension for studying language development in autistic children. Longitudinal monitoring of receptive-expressive balance captures within-child changes over time, revealing nuanced developmental processes that are not observable in cross-sectional studies. The heterogeneity observed across profiles, especially within ExpAdv, further suggests that receptive-expressive organization and overall language level represent related but distinct developmental dimensions. Examining these dimensions together may therefore provide a more comprehensive characterization of language variability in autism and help identify developmental patterns that may be obscured when language is considered only in terms of overall level.

### Clinical Implications

Our findings highlight receptive-expressive balance as a complementary dimension for characterizing language development in autistic children. Importantly, assessment of overall language level remains the primary consideration when evaluating whether a child has a language delay. Language delay can be assessed using different metrics (e.g., standardized scores, developmental quotients, developmental ages). Once the overall language level has been established, examining receptive and expressive abilities separately may provide additional information about relative strengths and vulnerabilities that can be overlooked when relying only on composite scores.

Language-specific assessments, such as the Preschool Language Scale (PLS-5; Zimmerman et al., 2011), provide detailed measures of receptive and expressive language and may be particularly informative when a more comprehensive language evaluation is possible. However, these assessments require more time and specialized training and may not always be feasible in routine clinical settings. Broader developmental assessments, such as the Mullen Scales of Early Learning (MSEL; Mullen, 1995), the Psychoeducational Profile–Third Edition (PEP-3; Schopler et al., 2005), or the Bayley Scales of Infant and Toddler Development (Bayley, 2012), represent practical alternatives, provided that receptive and expressive domains are interpreted separately rather than relying solely on composite scores.

Within this framework, an ExpAdv profile, indicating relatively stronger expressive than receptive abilities, may provide clinically relevant information. An ExpAdv profile does not necessarily reflect strong expressive language skills in absolute terms. When ExpAdv co-occurs with language delay, clinicians may consider closer longitudinal monitoring of both receptive and expressive language trajectories. In addition, relatively stronger expressive abilities may mask underlying comprehension difficulties, particularly when clinical impressions rely heavily on spoken language milestones. Early receptive abilities have also been shown to contribute to subsequent expressive language development in late-talker populations (Fisher, 2017).

Importantly, the clinical interpretation of receptive-expressive profiles remains preliminary and requires replication in independent longitudinal samples. These profiles should be viewed as developmental patterns that provide additional information beyond overall language level, rather than as diagnostic markers or definitive predictors of outcome. In our cohort, ExpAdv was not necessarily persistent, and many children subsequently transitioned toward Balanced or Receptive Advantage profiles. Nevertheless, the ExpAdv profile may be clinically informative when it occurs in the context of language delay, as children with ExpAdv showed greater social-communication difficulties and more frequent stereotyped and echolalic language despite relatively stronger expressive than receptive language scores. Thus, an apparent expressive advantage should not necessarily be interpreted as reflecting more generative language use or stronger functional communication and may instead signal a developmental pattern that warrants closer longitudinal monitoring.

Overall, while language level remains the primary basis for determining whether a child has a delay, receptive-expressive balance may provide a useful, more fine-grained characterization of language development and help highlight relative vulnerabilities warranting closer longitudinal attention. These findings provide an initial framework for clinical consideration, but further research is needed to establish their clinical and prognostic significance and to inform specific clinical recommendations.

### Limitations

Several limitations should be acknowledged. First, language was assessed using a single instrument (MSEL) that does not examine specific language domains (e.g., semantics, phonology, pragmatics). Moreover, although we applied cut-offs derived from a large TD sample in a previous study (Reinhartsen et al., 2019), the age range of our TD group was somewhat younger than that of the reference sample. The generalizability of these cut-offs across a wider age range and different language assessments remains to be established. Second, while the use of age-equivalent (AE) scores addressed floor effects associated with standard scores, AE scores have limitations. The receptive and expressive language AE scales do not have equal intervals, meaning that equivalent differences in AE scores may represent different amounts of developmental change across ages. In addition, ratio-based classifications may be less stable for children with scores near the floor on one scale, and profile membership may vary depending on the selected ratio cut-offs. Third, although the TD and autistic groups had similar age distributions at baseline, the samples differed in sex distribution and parental education. The potential influence of these factors was assessed by repeating all direct group-comparison analyses in a subsample matched on the joint distribution of sex and parental education, across 10 independent random draws. Results were largely consistent with those obtained in the full sample. Fourth, although the sliding-window approach provided fine-grained visualization of developmental changes, adjacent age windows were not statistically independent because individual observations could contribute to multiple overlapping windows (e.g., between 18–24 and 19–25 months). Consequently, age-related variation in profile prevalence should be interpreted as descriptive trends rather than independent estimates at each age window. Fifth, although the overall sample size was robust, subgroup analyses (e.g., comparisons between ExpAdv-Stable and ExpAdv-Switch) were limited by small sample sizes (N <15 per group in most windows), reducing statistical power. Notably, the significant sex-distribution contrast at the 30– 36 to 36–42-month transition involved a particularly small cell (one female among 29 ExpAdv-Switch children), and this result should be interpreted cautiously. Finally, follow-up into school age is needed to determine whether profile transitions continue and how they relate to later developmental outcomes.

### Conclusion

In this longitudinal study of 426 children (1,468 time points), we identified distinct receptive-expressive language trajectories and profiles in autism and TD. ExpAdv emerged as a relatively frequent profile in younger autistic children, associated with slower language and cognitive development, and greater social communication difficulties. In contrast, Balanced and RecAdv profiles showed moderate stability and reflected a continuum of more typical receptive-expressive organization, linked to more favorable developmental outcomes. Transition patterns indicated that ExpAdv is a profile that autistic children are more likely to move out of than into, suggesting its transient nature. These findings emphasize the importance of longitudinal monitoring of receptive and expressive skills to capture meaningful developmental changes and identify children who may experience less optimal language outcomes. Our supplementary analyses identified an exploratory association between ESDM exposure and transitions toward more balanced receptive-expressive profiles; however, this finding did not survive correction for multiple comparisons and should not be interpreted as evidence of an intervention effect.

## Acknowledgments section

The authors would like to thank all the families who participated in the study, as well as the many collaborators who contributed to data collection over the years.

## Authors contribution

Conceptualization, K.L., and M.S.; methodology, K.L., M.G., F.J., N.K., and M.S.; formal analysis, K.L.; writing—original draft preparation, K.L.; writing— review and editing, K.L., M.G., F.J., N.K., and M.S.; supervision, M.S.; funding acquisition, M.S. All authors have read and agreed to the published version of the manuscript.

## Ethical approval and informed consent statements

Written informed consent forms were signed and provided by the participants’ caregivers. The Ethics Committee of the University of Geneva approved the research protocol.

## Data availability statement

The datasets analyzed during the current study are not publicly available due to privacy and ethical restrictions but are available from the corresponding author upon reasonable request.

## Declaration of AI-Assisted Technologies

During the preparation of this manuscript, the authors used ChatGPT (OpenAI) and Claude (Anthropic) to assist with language editing and to support the generation of a Sankey diagram based exclusively on author-provided tabulated data. No AI tool was used to generate scientific results, perform data analyses, interpret findings, or draw conclusions. All outputs were reviewed, verified, and edited by the authors, who take full responsibility for the content of the manuscript and figures.

