## Supplementary material for "Longitudinal Receptive-Expressive Language Profiles in Young Autistic Children": -

**Table S1.** Sample characteristics of the TD and ASD groups matched on sex and parental education, summarized across 10 independent stratified random draws. ASD values are given as the range of draw-level means (mean SD across draws in parentheses); TD values are identical across draws by construction (fixed TD pool). p-values from χ2 (categorical) or Welch's t-test (continuous), computed separately per draw.

| Measure | Matched ASD (range across 10 draws) | Matched TD | p-value (range across draws) |
|---|---|---|---|
| **Number of participants** | 77 (all draws) | 77 (all draws) | — |
| **Number of time points** | 286–308 | 233 | — |
| **Female sex, n (%)** | 30 (39.0%) (all draws) | 30 (39.0%) (all draws) | 1.000 (χ2) |
| **Parental college education, n (%)** | 72 (93.5%) (all draws) | 72 (93.5%) (all draws) | 1.000 (χ2) |
| **Age at first visit, mean (SD)** | 2.6–2.9 (mean SD≈0.9) | 2.6 (1.1) | 0.128–0.938 [0/10 significant] |
| **ADOS CSS Total** | 7.6–8.0 (mean SD≈1.9) | 1.1 (0.2) | <0.001 [10/10 significant] |
| **ADOS CSS Social Affect** | 6.6–7.1 (mean SD≈2.1) | 1.1 (0.4) | <0.001 [10/10 significant] |
| **ADOS CSS RRB** | 8.7–9.0 (mean SD≈1.5) | 2.4 (2.0) | <0.001 [10/10 significant] |
| **MSEL Total DQ** | 62.8–68.3 (mean SD≈22.7) | 112.1 (13.1) | <0.001 [10/10 significant] |
| **MSEL EL DQ** | 49.7–55.7 (mean SD≈26.4) | 100.1 (19.7) | <0.001 [10/10 significant] |
| **MSEL RL DQ** | 50.0–55.2 (mean SD≈30.8) | 117.0 (17.3) | <0.001 [10/10 significant] |
| **MSEL VR DQ** | 76.1–83.6 (mean SD≈25.8) | 125.6 (23.3) | <0.001 [10/10 significant] |
| **MSEL FM DQ** | 73.3–79.3 (mean SD≈20.0) | 105.7 (16.3) | <0.001 [10/10 significant] |
| **VABS-II Adaptive Standard Score** | 77.3–80.5 (mean SD≈11.4) | 103.0 (7.8) | <0.001 [10/10 significant] |

**Table S2.** Results of the mixed model analysis comparing expressive and receptive linguistic skills between the TD and ASD groups. Note: P-value group effect evaluates the difference between mean scores in the two groups whereas p-value interaction refers to the difference in the shape of the developmental trajectory. The significance threshold was fixed at 0.05, and statistically significant values are shown in bold.
TD = Typical development; ASD = Autism spectrum disorder; RL = Receptive Language; EL = Expressive Language; AE = Age Equivalent scores.

| **TD vs. ASD** | **Group effect** | | | | | **Interaction with age** | | | |
|---|---|---|---|---|---|---|---|---|---|
| | **Model order** | ***p*-value** | **TD intercept (± SD)** | **ASD intercept (± SD)** | **Log likelihood, df** | ***p*-value** | **TD age slope (± SD)** | **ASD age slope (± SD)** | **Log likelihood, df** |
| **Receptive Language AE** | cubic | **<0.001** | 2.02 ± 5.20 | -3.59 ± 7.83 | 297.27, 4 | **<0.001** | -0.39 ± 0.40 | 1.33 ± 0.65 | 20.66, 3 |
| **Expressive Language AE** | cubic | **<0.001** | 5.14 ± 4.68 | 1.00 ± 7.07 | 300.65, 4 | **<0.001** | -0.49 ± 0.36 | 0.22 ± 0.59 | 97.30, 3 |
| **Within ASD** | **Model order** | ***p*-value** | **RL intercept (± SD)** | **EL intercept (± SD)** | **Log likelihood, df** | ***p*-value** | **RL age slope (± SD)** | **EL age slope (± SD)** | **Log likelihood, df** |
| **Receptive Language AE vs. Expressive Language AE** | quadratic | **<0.001** | -19.30 ± 2.07 | -24.86 ± 2.07 | 174.97, 3 | **< .001** | 1.22 ± 0.10 | 1.50 ± 0.10 | 36.50, 2 |
| **Within TD** | **Model order** | ***p*-value** | **RL intercept (± SD)** | **EL intercept (± SD)** | **Log likelihood, df** | ***p*-value** | **RL age slope (± SD)** | **EL age slope (± SD)** | **Log likelihood, df** |
| **Receptive Language AE vs. Expressive Language AE** | quadratic | **<0.001** | -23.56 ± 2.55 | -11.27 ± 2.55 | 78.84, 3 | **.002** | 2.38 ± 0.14 | 1.98 ± 0.14 | 12.86, 2 |

**Table S3.** Developmental trajectories of expressive (EL) and receptive (RL) language age-equivalents (MSEL), ASD vs. TD, across 10 independent stratified random draws (matched on sex and parental education). Group effect = main effect of Group (ASD vs TD) on DQ/age-equivalent. Group×Age interaction = whether trajectories diverge over age. Yellow cells to be filled in after running the **mixed-model script on each draw.**

| | | | Expressive Language (EL) | | | Receptive Language (RL) | | |
|---|---|---|---|---|---|---|---|---|
| **Draw** | **N ASD visits** | **N TD visits** | **Group effect p-value** | **Group × Age interaction p-value** | **Direction of effect** | **Group effect p-value** | **Group×Age interaction p-value** | **Direction of effect** |
| **1** | 289 | 233 | <0.001 | <0.001 | ASD: lower and slower EL development | <0.001 | 0.121 | ASD: lower RL development |
| **2** | 290 | 233 | <0.001 | <0.001 | ASD: lower and slower EL development | <0.001 | 0.024 | ASD: lower and slower RL development |
| **3** | 306 | 233 | <0.001 | <0.001 | ASD: lower and slower EL development | <0.001 | 0.115 | ASD: lower RL development |
| **4** | 295 | 233 | <0.001 | <0.001 | ASD: lower and slower EL development | <0.001 | 0.003 | ASD: lower and slower RL development |
| **5** | 306 | 233 | <0.001 | <0.001 | ASD: lower and slower EL development | <0.001 | 0.012 | ASD: lower and slower RL development |
| **6** | 306 | 233 | <0.001 | <0.001 | ASD: lower and slower EL development | <0.001 | 0.047 | ASD: lower and slower RL development |
| **7** | 308 | 233 | <0.001 | <0.001 | ASD: lower and slower EL development | <0.001 | 0.139 | ASD: lower RL development |
| **8** | 308 | 233 | <0.001 | <0.001 | ASD: lower and slower EL development | <0.001 | 0.023 | ASD: lower and slower RL development |
| **9** | 286 | 233 | <0.001 | <0.001 | ASD: lower and slower EL development | <0.001 | 0.001 | ASD: lower and slower RL development |
| **10** | 292 | 233 | <0.001 | <0.001 | ASD: lower and slower EL development | <0.001 | 0.001 | ASD: lower and slower RL development |
| **Significant in** | | | **10/10 draws** | **10/10 draws** | | **10/10 draws** | **7/10 draws** | |

Across 10 independent stratified random draws of the sex- and parental-education-matched subsample, the expressive language (EL) group difference was fully robust: autistic children showed significantly lower EL scores than TD peers in all 10 draws ($p < 0.001$), and this gap significantly widened with age (Group×Age interaction, $p < 0.001$ in 10/10 draws). This indicates consistently divergent EL trajectories regardless of which children were sampled into the matched ASD group. The receptive language (RL) group difference was robust for the main effect but not for the interaction: autistic children showed significantly lower RL scores than TD peers in all 10 draws ($p < 0.001$), but the Group×Age interaction — reflecting whether the ASD–TD gap widens over age — reached significance in only 7/10 draws (range: $p = 0.001$–$0.139$). In the 3 draws where it did not reach significance, the effect was in the same direction but did not cross the significance threshold.

**Table S4.** Distribution of language profiles (Expressive Advantage, Balanced, Receptive Advantage) in the autistic and typically developing (TD) groups. restricted to visits between 18 and 54 months (ASD: n=783 visits; TD: n=203 visits). Group differences were assessed using a multinomial logistic regression with age-window as covariate. Odd ratios (ORs) reflect the odds of ASD relative to TD for each profile versus Balanced (reference category). LR: likelihood ratio.

| Profile | ASD visits | % | TD visits | % | P-value | OR |
|---|---|---|---|---|---|---|
| **ExpAdv** | 200 | 26% | 20 | 10% | **<0.001** | 3.61 [2.13–6.13] |
| **Balanced** | 325 | 41% | 97 | 48% | **0.013** | 0.66 [0.48–0.92] |
| **RecAdv** | 258 | 33% | 86 | 42% | 0.844 | 1.04 [0.73–1.47] |
| **Total** | 783 | 100% | 203 | 100% | 0.006 | LR $\chi^2(2) = 30.87$ |

Overall group differences in profile distribution were assessed using a multinomial logistic regression controlling for age-window (LR $\chi^2(2) = 30.87$, $p$ <0.001). The ExpAdv profile was significantly more frequent in ASD than TD ($p$ <0.001), Balanced profiles were significantly more frequent in TD than ASD ($p$ = 0.013), and RecAdv proportions did not differ significantly between groups (p = 0.844)."

**Table S5.** Language-profile distribution (ExpAdv, Balanced, RecAdv) in ASD vs. TD visits, 18–54 months, across 10 independent stratified random draws (matched on sex and parental education). Multinomial logistic regression with continuous age (months) as covariate. ORs (ASD vs. TD, age-adjusted) from per-profile binary logistic regressions (profile vs. other two combined).

Across 10 independent stratified random draws of the sex- and parental-education-matched subsample, the ExpAdv profile remained significantly more frequent in ASD than TD in 9/10 draws (OR range: 1.56–2.48), Balanced profiles were significantly more frequent in TD in 5/10 draws (OR range: 0.60–0.76), and RecAdv proportions never differed significantly between groups (0/10 draws; OR range: 0.81–1.16). The overall group effect (LR $\chi^2$) was significant in 9/10 draws. The direction and general pattern of effects were therefore stable across random draws, though statistical significance for the Balanced profile specifically was sensitive to the particular subsample drawn.

| Draw | N ASD visits | N TD visits | ExpAdv OR [95% CI] | ExpAdv p | Balanced OR [95% CI] | Balanced p | RecAdv OR [95% CI] | RecAdv p | LR $\chi^2(2)$ | Overall p |
|---|---|---|---|---|---|---|---|---|---|---|
| 1 | 232 | 192 | 2.34 [1.34–4.08] | 0.003 | 0.60 [0.40–0.90] | 0.014 | 1.03 [0.68–1.55] | 0.894 | 11.29 | 0.004 |
| 2 | 241 | 192 | 2.03 [1.17–3.53] | 0.012 | 0.60 [0.40–0.89] | 0.011 | 1.14 [0.76–1.71] | 0.513 | 9.38 | 0.009 |
| 3 | 250 | 192 | 2.44 [1.42–4.20] | 0.001 | 0.70 [0.48–1.04] | 0.078 | 0.85 [0.57–1.27] | 0.425 | 11.24 | 0.004 |
| 4 | 237 | 192 | 2.48 [1.44–4.27] | 0.001 | 0.60 [0.40–0.89] | 0.012 | 0.98 [0.65–1.47] | 0.928 | 12.97 | 0.002 |
| 5 | 237 | 192 | 1.56 [0.88–2.77] | 0.130 | 0.70 [0.47–1.04] | 0.080 | 1.16 [0.77–1.73] | 0.483 | 3.96 | 0.138 |
| 6 | 247 | 192 | 2.20 [1.27–3.81] | 0.005 | 0.67 [0.45–0.99] | 0.044 | 0.97 [0.64–1.45] | 0.867 | 9.21 | 0.010 |
| 7 | 256 | 192 | 2.32 [1.35–3.98] | 0.002 | 0.73 [0.49–1.08] | 0.112 | 0.85 [0.57–1.26] | 0.415 | 9.92 | 0.007 |
| 8 | 253 | 192 | 1.90 [1.10–3.30] | 0.022 | 0.67 [0.45–0.99] | 0.043 | 1.07 [0.72–1.59] | 0.745 | 6.91 | 0.032 |
| 9 | 223 | 192 | 2.44 [1.40–4.24] | 0.002 | 0.69 [0.46–1.04] | 0.077 | 0.85 [0.56–1.30] | 0.457 | 10.67 | 0.005 |
| 10 | 237 | 192 | 2.31 [1.33–4.01] | 0.003 | 0.76 [0.51–1.13] | 0.175 | 0.81 [0.54–1.23] | 0.323 | 9.35 | 0.009 |
| **Significant in** | | | | **9/10 draws** | | **5/10 draws** | | **0/10 draws** | | **9/10 draws** |
| **OR range** | | | **1.56–2.48** | | **0.60–0.76** | | **0.81–1.16** | | | |

**Table S6.** Profile distribution across age windows in the autistic group. N and percentage of children classified as Expressive Advantage, Receptive Advantage, or Balanced at each 6-month sliding window from 18 to 54 months.

| Age Window | N ExpAdv | N RecAdv | N Balanced | N Total | % ExpAdv | % RecAdv | % Balanced |
|---|---|---|---|---|---|---|---|
| 18-24 mo | 16 | 12 | 19 | 47 | 34.0% | 25.5% | 40.4% |
| 19-25 mo | 21 | 18 | 20 | 59 | 35.6% | 30.5% | 33.9% |
| 20-26 mo | 27 | 21 | 20 | 68 | 39.7% | 30.9% | 29.4% |
| 21-27 mo | 30 | 26 | 23 | 79 | 38.0% | 32.9% | 29.1% |
| 22-28 mo | 34 | 28 | 31 | 93 | 36.6% | 30.1% | 33.3% |
| 23-29 mo | 38 | 31 | 34 | 103 | 36.9% | 30.1% | 33.0% |
| 24-30 mo | 42 | 32 | 40 | 114 | 36.8% | 28.1% | 35.1% |
| 25-31 mo | 51 | 34 | 43 | 128 | 39.8% | 26.6% | 33.6% |
| 26-32 mo | 51 | 41 | 56 | 148 | 34.5% | 27.7% | 37.8% |
| 27-33 mo | 53 | 45 | 58 | 156 | 34.0% | 28.8% | 37.2% |
| 28-34 mo | 53 | 50 | 60 | 163 | 32.5% | 30.7% | 36.8% |
| 29-35 mo | 55 | 57 | 67 | 179 | 30.7% | 31.8% | 37.4% |
| 30-36 mo | 57 | 68 | 63 | 188 | 30.3% | 36.2% | 33.5% |
| 31-37 mo | 51 | 72 | 69 | 192 | 26.6% | 37.5% | 35.9% |
| 32-38 mo | 58 | 75 | 68 | 201 | 28.9% | 37.3% | 33.8% |
| 33-39 mo | 58 | 73 | 79 | 210 | 27.6% | 34.8% | 37.6% |
| 34-40 mo | 54 | 80 | 80 | 214 | 25.2% | 37.4% | 37.4% |
| 35-41 mo | 50 | 84 | 82 | 216 | 23.1% | 38.9% | 38.0% |
| 36-42 mo | 46 | 75 | 85 | 206 | 22.3% | 36.4% | 41.3% |
| 37-43 mo | 45 | 75 | 90 | 210 | 21.4% | 35.7% | 42.9% |
| 38-44 mo | 40 | 70 | 91 | 201 | 19.9% | 34.8% | 45.3% |
| 39-45 mo | 37 | 64 | 100 | 201 | 18.4% | 31.8% | 49.8% |
| 40-46 mo | 37 | 55 | 95 | 187 | 19.8% | 29.4% | 50.8% |
| 41-47 mo | 35 | 57 | 94 | 186 | 18.8% | 30.6% | 50.5% |
| 42-48 mo | 30 | 60 | 106 | 196 | 15.3% | 30.6% | 54.1% |
| 43-49 mo | 25 | 64 | 102 | 191 | 13.1% | 33.5% | 53.4% |
| 44-50 mo | 25 | 69 | 98 | 192 | 13.0% | 35.9% | 51.0% |
| 45-51 mo | 24 | 73 | 91 | 188 | 12.8% | 38.8% | 48.4% |
| 46-52 mo | 25 | 78 | 92 | 195 | 12.8% | 40.0% | 47.2% |
| 47-53 mo | 23 | 74 | 84 | 181 | 12.7% | 40.9% | 46.4% |
| 48-54 mo | 22 | 68 | 84 | 174 | 12.6% | 39.1% | 48.3% |

**Table S7.** Profile distribution across age windows in the typically developing group. N and percentage of children classified as Expressive Advantage, Receptive Advantage, or Balanced at each 6-month sliding window from 18 to 54 months.

| Age Window | N ExpAdv | N RecAdv | N Balanced | N Total | % ExpAdv | % RecAdv | % Balanced |
|---|---|---|---|---|---|---|---|
| 18-24 mo | 1 | 28 | 5 | 34 | 2.9% | 82.4% | 14.7% |
| 19-25 mo | 1 | 27 | 8 | 36 | 2.8% | 75.0% | 22.2% |
| 20-26 mo | 1 | 30 | 9 | 40 | 2.5% | 75.0% | 22.5% |
| 21-27 mo | 2 | 27 | 11 | 40 | 5.0% | 67.5% | 27.5% |
| 22-28 mo | 3 | 24 | 14 | 41 | 7.3% | 58.5% | 34.1% |
| 23-29 mo | 4 | 28 | 18 | 50 | 8.0% | 56.0% | 36.0% |
| 24-30 mo | 4 | 29 | 21 | 54 | 7.4% | 53.7% | 38.9% |
| 25-31 mo | 4 | 24 | 20 | 48 | 8.3% | 50.0% | 41.7% |
| 26-32 mo | 3 | 19 | 24 | 46 | 6.5% | 41.3% | 52.2% |
| 27-33 mo | 3 | 19 | 27 | 49 | 6.1% | 38.8% | 55.1% |
| 28-34 mo | 4 | 17 | 25 | 46 | 8.7% | 37.0% | 54.3% |
| 29-35 mo | 4 | 12 | 25 | 41 | 9.8% | 29.3% | 61.0% |
| 30-36 mo | 4 | 12 | 24 | 40 | 10.0% | 30.0% | 60.0% |
| 31-37 mo | 5 | 10 | 26 | 41 | 12.2% | 24.4% | 63.4% |
| 32-38 mo | 6 | 11 | 24 | 41 | 14.6% | 26.8% | 58.5% |
| 33-39 mo | 5 | 10 | 21 | 36 | 13.9% | 27.8% | 58.3% |
| 34-40 mo | 3 | 9 | 24 | 36 | 8.3% | 25.0% | 66.7% |
| 35-41 mo | 4 | 10 | 22 | 36 | 11.1% | 27.8% | 61.1% |
| 36-42 mo | 4 | 7 | 20 | 31 | 12.9% | 22.6% | 64.5% |
| 37-43 mo | 5 | 7 | 22 | 34 | 14.7% | 20.6% | 64.7% |
| 38-44 mo | 5 | 7 | 21 | 33 | 15.2% | 21.2% | 63.6% |
| 39-45 mo | 5 | 5 | 21 | 31 | 16.1% | 16.1% | 67.7% |
| 40-46 mo | 7 | 7 | 23 | 37 | 18.9% | 18.9% | 62.2% |
| 41-47 mo | 6 | 7 | 23 | 36 | 16.7% | 19.4% | 63.9% |
| 42-48 mo | 6 | 8 | 26 | 40 | 15.0% | 20.0% | 65.0% |
| 43-49 mo | 6 | 8 | 29 | 43 | 14.0% | 18.6% | 67.4% |
| 44-50 mo | 5 | 11 | 28 | 44 | 11.4% | 25.0% | 63.6% |
| 45-51 mo | 7 | 11 | 26 | 44 | 15.9% | 25.0% | 59.1% |
| 46-52 mo | 5 | 8 | 23 | 36 | 13.9% | 22.2% | 63.9% |
| 47-53 mo | 5 | 7 | 25 | 37 | 13.5% | 18.9% | 67.6% |
| 48-54 mo | 6 | 6 | 24 | 36 | 16.7% | 16.7% | 66.7% |

**Table S8.** Transition probabilities between language profiles across consecutive age windows for the autistic group (18-54 months). ExpAdv = Expressive Advantage; Balanced = Balanced; RecAdv = Receptive Advantage. Diagonal values indicate stability within the same language profile.

| Transition | From → | ExpAdv | Balanced | RecAdv |
|---|---|---|---|---|
| **18-24 to 24-30 (n=39)** | **ExpAdv (n=12)** | **50.0%** | 33.3% | 16.7% |
| | **Balanced (n=18)** | 22.2% | **50.0%** | 27.8% |
| | **RecAdv (n=9)** | 0.0% | 33.3% | **66.7%** |
| **24-30 to 30-36 (n=88)** | **ExpAdv (n=29)** | **48.3%** | 31.0% | 20.7% |
| | **Balanced (n=31)** | 12.9% | **51.6%** | 35.5% |
| | **RecAdv (n=28)** | 7.1% | 33.3% | **60.7%** |
| **30-36 to 36-42 (n=145)** | **ExpAdv (n=47)** | **38.3%** | 40.4% | 21.3% |
| | **Balanced (n=49)** | 22.5% | **34.7%** | 42.9% |
| | **RecAdv (n=49)** | 6.1% | 49.0% | **44.9%** |
| **36-42 to 42-48 (n=154)** | **ExpAdv (n=35)** | **22.9%** | 57.1% | 20.0% |
| | **Balanced (n=63)** | 12.7% | **63.5%** | 23.8% |
| | **RecAdv (n=56)** | 1.8% | 46.4% | **51.8%** |
| **42-48 to 48-54 (n=137)** | **ExpAdv (n=22)** | **40.9%** | 40.9% | 18.2% |
| | **Balanced (n=75)** | 2.7% | **60.0%** | 37.3% |
| | **RecAdv (n=40)** | 7.5% | 35.0% | **57.5%** |

**Table S9.** Baseline characteristics and adaptive behavior scores at each developmental transition, comparing children who remained in the ExpAdv profile (ExpAdv-Stable) with those who transitioned to a Balanced or RecAdv profile (ExpAdv-Switch).
For each consecutive age-window transition (18–24 to 24–30, 24–30 to 30–36, 30–36 to 36–42, 36–42 to 42–48, and 42–48 to 48–54 months), group differences in sex ratio, ESDM intervention proportion, and VABS-II standard scores (Adaptive Behavior, Communication, Socialization, and Daily Living Skills) were compared between groups. Categorical variables were analyzed using Fisher's Exact Test (FET) and continuous variables using independent samples t-tests, with Cohen's *d* reported as a measure of effect size. Bold P-values indicate statistical significance ($p < 0.05$). P-values were adjusted for multiple comparisons using the Bonferroni correction. Bold P-values indicate statistical significance after Bonferroni correction (adjusted $p < 0.0083$).

| **Transition** | | **ExpAdv-Stable (n = 6)** | **ExpAdv-Switch (n = 6)** | ***P*-value** | **Cohen's *d*** |
|---|---|---|---|---|---|
| **18–24 to 24–30 months** | N Females (%) | 6 (100%) | 3 (50%) | 0.182 (FET) | |
| | % ESDM Intervention | 4 (66.7%) | 4 (66.7%) | 1.000 (FET) | |
| | Mean Adaptive Standard Score (SD) (VABS-II) | 79.50 (6.83) | 77.67 (8.24) | 0.684 | 0.242 |
| | Mean Communication Standard Score (SD) (VABS-II) | 71.50 (4.28) | 66.33 (6.62) | 0.340 | 0.927 |
| | Mean Socialization Standard Score (SD) (VABS-II) | 82.00 (4.00) | 83.33 (7.97) | 0.946 | -0.212 |
| | Mean Daily Living Skills Standard Score (SD) (VABS-II) | 84.00 (9.06) | 81.17 (8.35) | 0.586 | 0.325 |
| **Transition** | | **ExpAdv-Stable** (n = 14) | **ExpAdv-Switch** (n = 15) | ***P*-value** | **Cohen's *d*** |
| **24–30 to 30–36 months** | N Females (%) | 1 (7.14%) | 2 (13.33%) | 1.000 (FET) | |
| | % ESDM Intervention | 9 (64.3%) | 14 (93.3%) | 0.080 (FET) | |
| | Mean Adaptive Standard Score (SD) (VABS-II) | 81.00 (8.10) | 80.00 (8.71) | 0.752 | 0.119 |
| | Mean Communication Standard Score (SD) (VABS-II) | 75.64 (9.44) | 74.47 (11.31) | 0.442 | 0.113 |
| | Mean Socialization Standard Score (VABS-II) | 82.93 (7.09) | 82.00 (9.29) | 0.582 | 0.112 |
| | Mean Daily Living Skills Standard Score (SD) (VABS-II) | 81.71 (11.67) | 85.67 (9.71) | 0.196 | -0.369 |
| **Transition** | | **ExpAdv-Stable** (n = 18) | **ExpAdv-Switch** (n = 29) | ***P*-value** | **Cohen's *d*** |
| **30–36 to 36–42 months** | N Females (%) | 9 (50.0%) | 1 (3.45%) | **<0.001** (FET) | |
| | % ESDM Intervention | 9 (50.0%) | 25 (86.2%) | 0.017 (FET) | |
| | Mean Adaptive Standard Score (SD) (VABS-II) | 74.56 (8.02) | 78.31 (8.68) | 0.145 | -0.445 |
| | Mean Communication Standard Score (SD) (VABS-II) | 66.44 (6.19) | 75.31 (11.37) | **0.004** | -0.910 |
| | Mean Socialization Standard Score (VABS-II) | 75.67 (8.40) | 78.52 (7.89) | 0.246 | -0.352 |
| | Mean Daily Living Skills Standard Score (SD) (VABS-II) | 78.89 (10.51) | 82.62 (12.49) | 0.297 | -0.317 |
| **Transition** | | **ExpAdv-Stable** (n = 8) | **ExpAdv-Switch** (n =27) | ***P*-value** | **Cohen's *d*** |
| **36–42 to 42–48 months** | N Females (%) | 5 (62.5%) | 19 (70.4%) | 0.116 (FET) | |
| | % ESDM Intervention | 4 (50.0%) | 17 (63.0%) | 0.685 (FET) | |
| | Mean Adaptive Standard Score (SD) (VABS-II) | 71.25 (5.70) | 76.33 (10.36) | 0.196 | -0.531 |
| | Mean Communication Standard Score (SD) (VABS-II) | 64.38 (7.25) | 71.63 (15.54) | 0.221 | -0.511 |
| | Mean Socialization Standard Score (VABS-II) | 72.88 (5.49) | 76.74 (9.48) | 0.282 | -0.440 |
| | Mean Daily Living Skills Standard Score (SD) (VABS-II) | 77.50 (8.04) | 79.04 (11.63) | 0.730 | -0.140 |
| **Transition** | | **ExpAdv-Stable** (n = 9) | **ExpAdv-Switch** (n = 13) | ***P*-value** | **Cohen's *d*** |
| **42–48 to 48–54 months** | N Females (%) | 3 (33.3%) | 5 (38.5%) | 1.000 (FET) | |
| | % ESDM Intervention | 9 (50.0%) | 25 (86.2%) | 0.017 (FET) | |
| | Mean Adaptive Standard Score (SD) (VABS-II) | 71.44 (9.08) | 71.08 (8.59) | 0.924 | 0.042 |
| | Mean Communication Standard Score (SD) (VABS-II) | 66.78 (15.64) | 66.69 (13.82) | 0.989 | 0.006 |
| | Mean Socialization Standard Score (VABS-II) | 73.00 (8.92) | 74.92 (6.87) | 0.574 | -0.248 |
| | Mean Daily Living Skills Standard Score (SD) (VABS-II) | 78.56 (10.91) | 74.62 (9.84) | 0.387 | 0.383 |

**Table S10.** ADOS severity scores and item-level comparison scores ExpAdv, Balanced, and RecAdv profiles.
Median scores (Q1–Q3) for each ADOS-2 item are presented for the Expressive Advantage (ExpAdv), Balanced, and Receptive Advantage (RecAdv) profiles. Kruskal-Wallis H statistics and p-values are reported for overall group comparisons. Mann-Whitney pairwise comparisons between ExpAdv vs. Balanced, ExpAdv vs. RecAdv, and Balanced vs. RecAdv are conducted if the Kruskal-Wallis tests are significant. Items are grouped by domain: Calibrated Severity Score, Social Communication, Social Interaction, and Repetitive and Restricted Behaviors. Significant p-values ($p < .05$) are highlighted in bold. *P*-values of Mann-Whitney comparisons are corrected with Bonferroni ($p = 0.0167$).

| Category | Item | ExpAdv Median (Q1-Q3) | Balanced Median (Q1-Q3) | RecAdv Median (Q1-Q3) | Kruskal-Wallis H | *P*-value | ExpAdv *vs.* Balanced U | *P*-value | ExpAdv *vs.* RecAdv U | *P*-value | *Balanced vs. RecAdv U* | *P*-value |
|---|---|---|---|---|---|---|---|---|---|---|---|---|
| **Calibrated Severity Score** | Total Score | 9 (7-10) | 7 (6-9) | 7 (6-9) | 14.76 | **<0.001** | 3949 | **<0.001** | 3775 | **<0.001** | 4555 | 0.894 |
| | Social Affect Score | 8 (6-9) | 6 (4-8) | 6 (5-8) | 14.33 | **<0.001** | 3797 | **<0.001** | 4027 | 0.008 | 4250 | 0.351 |
| | RRB Score | 10 (8.5-10) | 10 (8.75-10) | 9 (7.75-10) | 7.41 | **0.025** | 5154 | 0.623 | 4161 | **0.011** | 3864 | 0.037 |
| **Social Communication** | Gestures: Descriptive, Instrumental, Informational | 1 (1-2) | 1 (0-1.25) | 1 (0-1) | 9.52 | **0.009** | 4219 | **0.007** | 4061 | 0.009 | 4580 | 0.942 |
| | Immediate Echolalia | 2 (1-2) | 1 (1-2) | 1 (0-2) | 7.79 | **0.020** | 3633 | **0.005** | 3488 | 0.057 | 4096 | 0.630 |
| | Intonation | 2 (1.5-2) | 2 (1-2) | 2 (1-2) | 4.42 | 0.110 | 3897 | 0.047 | 3180 | 0.112 | 3497 | 0.916 |
| | Pointing | 3 (1-3) | 1 (1-3) | 1 (1-3) | 11.71 | **0.003** | 4328 | 0.088 | 3503 | **<0.001** | 3685 | 0.138 |
| | Stereotyped Language | 2 (1-3) | 1 (1-2) | 1 (1-2) | 9.49 | **0.009** | 602 | **0.006** | 996 | 0.978 | 472 | **0.007** |
| | Use of Another's Body to Communicate | 0 (1-2) | 0 (0-1) | 1 (0-2) | 3.82 | 0.148 | | | | | | |
| | Frequency of Spontaneous Vocalizations Directed to Others | 2 (1-3) | 1 (0.5-2) | 2 (1-2) | 24.22 | **<0.001** | 2704 | **<0.001** | 3610 | 0.018 | 2822 | **0.010** |
| **Social Interaction** | Facial Expressions Directed to Others | 1 (1-2) | 1 (1-2) | 1 (1-2) | 5.57 | 0.0617 | | | | | | |
| | Giving | 1 (1-2) | 0 (1-1.5) | 1 (0-2) | 11.64 | **0.003** | 3216 | **<0.001** | 4055 | 0.311 | 2918 | 0.023 |
| | Initiation of Joint Attention | 1 (0-3) | 1 (0-2) | 1 (0-2) | 3.66 | 0.161 | | | | | | |
| | Integration of Gaze and Other Behaviors during Social Overtures | 2 (1-2) | 1 (1-2) | 1 (1-2) | 8.15 | 0.017 | 3474 | **0.004** | 3994 | 0.177 | 3218 | 0.177 |
| | Response to Name | 2 (0-3) | 1 (0-2) | 1 (0-2) | 21.83 | **<0.001** | 3193 | **<0.001** | 3193 | **<0.001** | 3199 | **<0.001** |
| | Quality of Rapport | 2 (2-3) | 1 (1-2) | 1 (1-2) | 30.58 | **<0.001** | 3224 | **<0.001** | 2955 | **<0.001** | 4227 | 0.889 |
| | Quality of Social Overtures | 2 (2-3) | 1.5 (1-2) | 2 (1-2) | 27.00 | **<0.001** | 3494 | **<0.001** | 3579 | **<0.001** | 4439 | 0.606 |
| | Quality of Social Response | 2 (2-2) | 1 (1-2) | 1 (1-2) | 14.67 | **<0.001** | 738 | **<0.001** | 720 | **<0.001** | 1282 | 0.881 |
| | Quantity of Social Overtures | 2 (2-3) | 1 (0-2) | 2 (0.75-3) | 22.52 | **<0.001** | 3123 | **<0.001** | 3486 | **<0.001** | 3735 | 0.194 |
| | Requesting | 1 (0-2) | 0 (0-1) | 0 (0-2) | 11.64 | **0.003** | 3238 | **<0.001** | 3587 | 0.025 | 3353 | 0.458 |
| | Response to Joint Attention | 1 (0-2) | 0 (0-1) | 0 (0-2) | 18.11 | **<0.001** | 3094 | **<0.001** | 3154 | **<0.001** | 3815 | 0.849 |
| | Shared Enjoyment in Interaction | 1 (0-2) | 0 (0-1) | 0 (0-1) | 12.33 | **0.002** | 4003 | **<0.001** | 4198 | 0.017 | 4340 | 0.432 |
| | Showing | 2 (1-3) | 1 (0-2) | 2 (0-2) | 11.12 | **0.004** | 3767 | **0.001** | 4011 | 0.035 | 3758 | 0.169 |
| **Repetitive and Restricted Behaviors** | Hand and Finger and other Complex Mannerisms | 3 (2-3) | 2 (1.75-3) | 2 (1-3) | 3.45 | 0.179 | 5049 | 0.464 | 4403 | 0.065 | 4213 | 0.280 |
| | Repetitive Interests and Behaviors | 3 (2-3) | 2 (2-3) | 2 (1-3) | 17.62 | **<0.001** | 4127 | **0.003** | 3517 | **<0.001** | 4199 | 0.264 |
| | Self-Injurious Behaviors | 0 (0-0) | 0 (0-0) | 0 (0-0) | 3.80 | 0.149 | | | | | | |
| | Unusual Sensory Interests | 3 (1-3) | 2 (1-3) | 2 (1-3) | 12.55 | **0.002** | 4296 | **0.010** | 3803 | **<0.001** | 4216 | 0.294 |

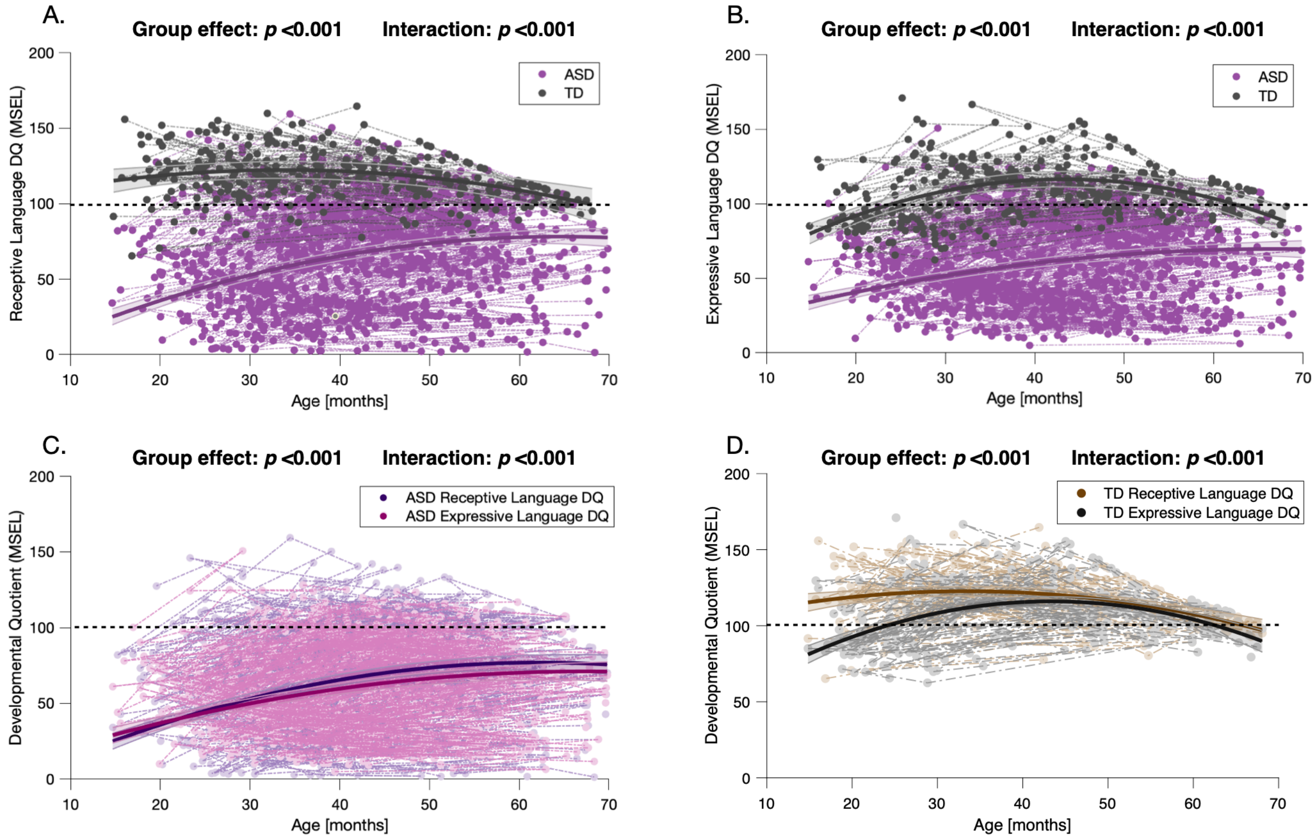


**Figure S1.** Trajectories of receptive and expressive language developmental quotients (MSEL) across age in the ASD and TD groups. (A) Receptive Language trajectories in ASD and TD participants. (B) Expressive Language trajectories in ASD and TD participants. (C) Overlay of receptive and expressive language trajectories within the ASD group. (D) Overlay of receptive and expressive language trajectories within the TD group. between the ASD and TD groups, as well as (C) within-ASD trajectories and (D) within-TD trajectories

The colored bands around the estimated group-level trajectory indicate the 95% confidence interval. MSEL : Mullen Scales of Early Learning ; ASD : Autism Spectrum Disorder ; TD: typical development ; DQ : Developmental Quotient.

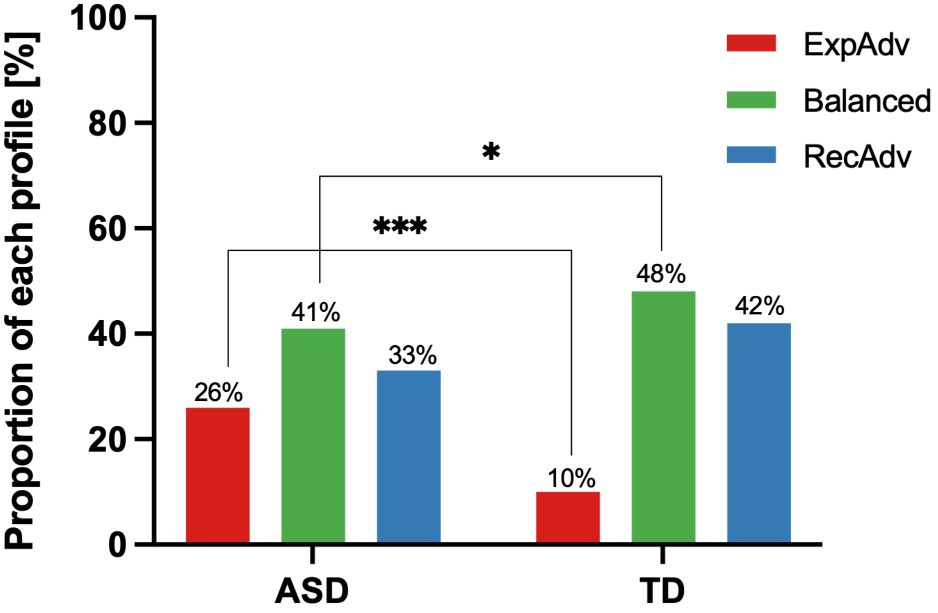


**Figure S2.** Proportions of Expressive Language, Balanced, and Receptive Language in the autistic and TD visits between 18 and 54 months (n= 783 visits from ASD; n=203 visits from TD). Proportions reflect age-adjusted marginal estimates from a multinomial logistic regression controlling for age-window. *$p$ <0.05 ***$p$ <0.001.

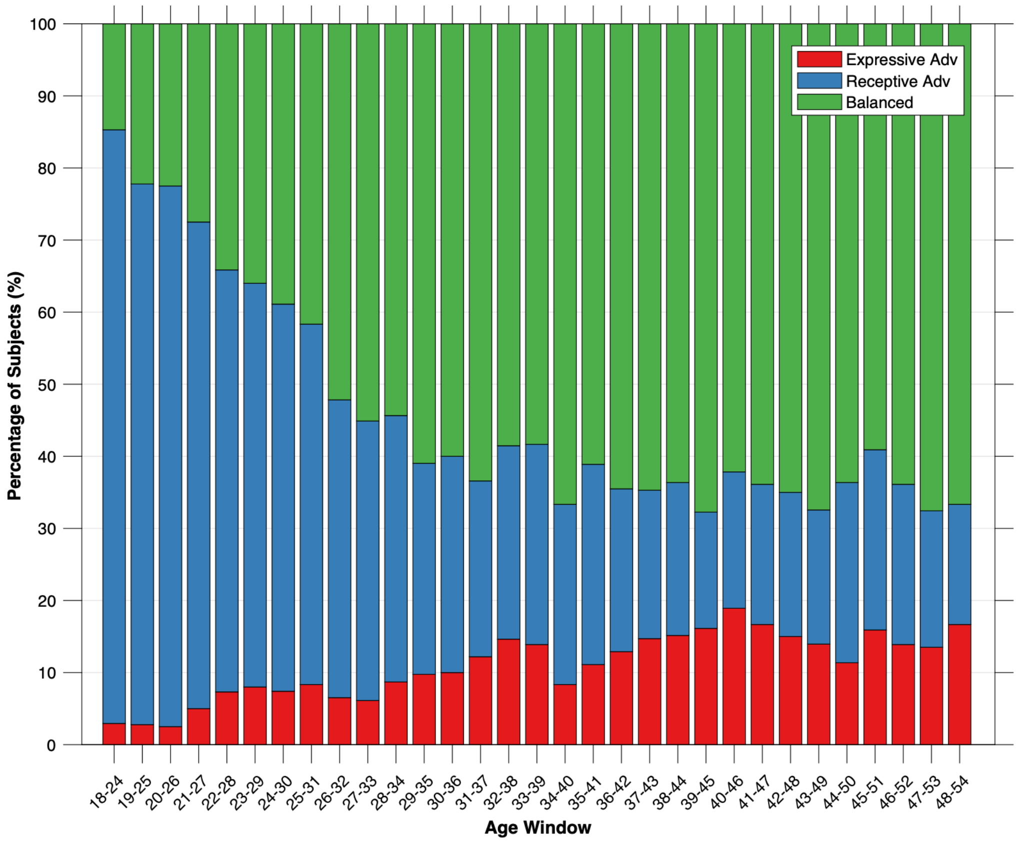


**Figure S3.** Proportion of Expressive Advantage, Balanced, and Receptive Advantage profiles within each age window in the TD group.

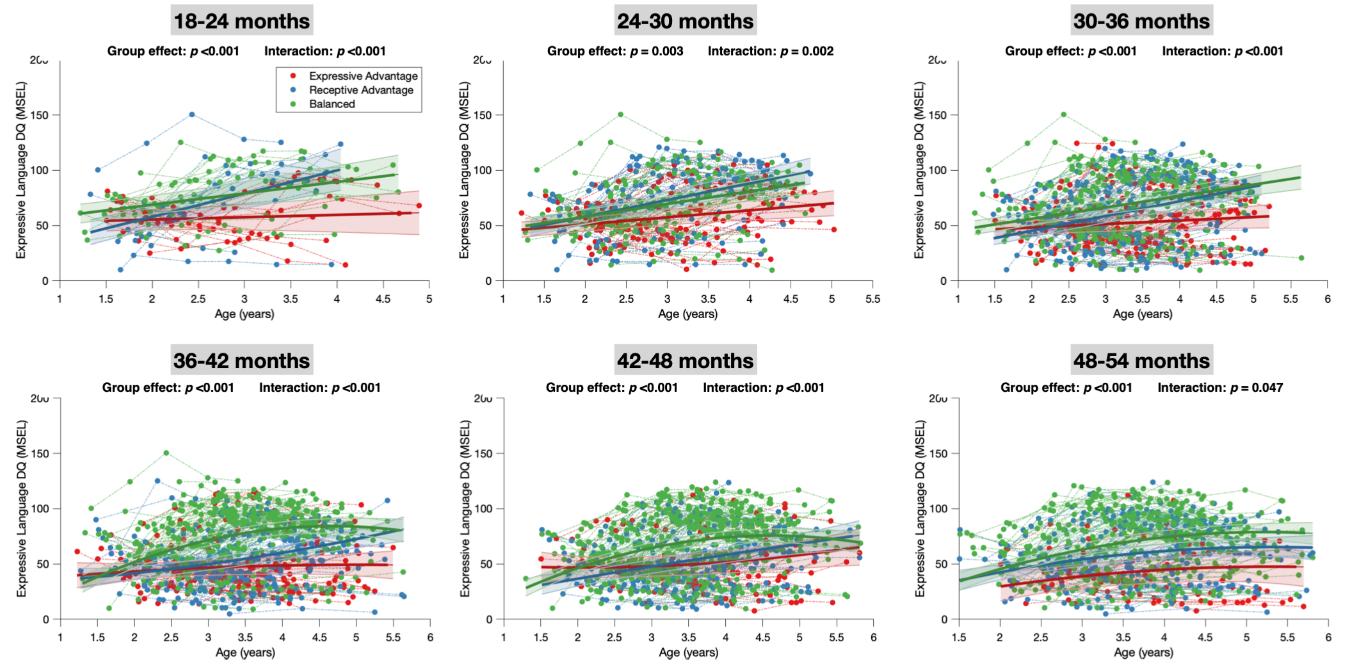


**Figure S4.** Trajectories of expressive language development quotient of the Expressive Advantage, Receptive Advantage, and Balanced profiles in the ASD group across six age windows.

The colored bands around the estimated group-level trajectory indicate the 95% confidence interval. MSEL : Mullen Scales of Early Learning ; DQ: Developmental Quotient.

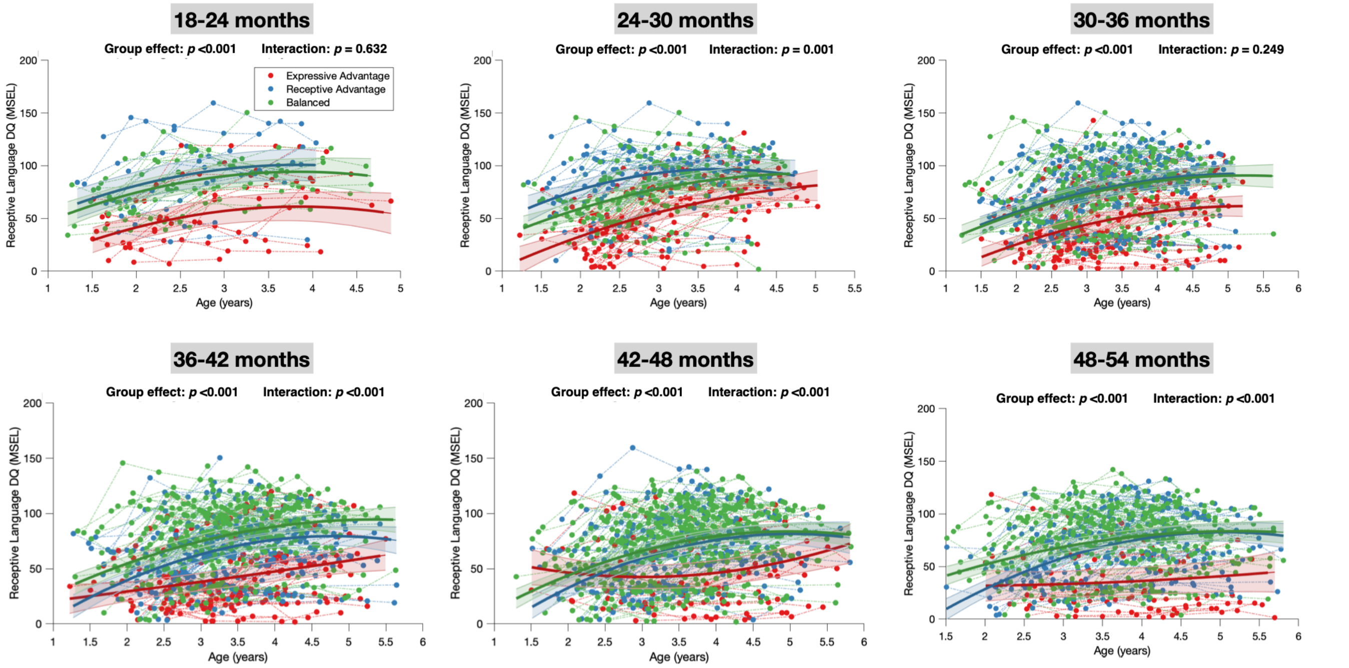


**Figure S5.** Trajectories of receptive language development quotient of the Expressive Advantage, Receptive Advantage, and Balanced profiles in the ASD group across six age windows.

The colored bands around the estimated group-level trajectory indicate the 95% confidence interval. MSEL : Mullen Scales of Early Learning ; DQ: Developmental Quotient.

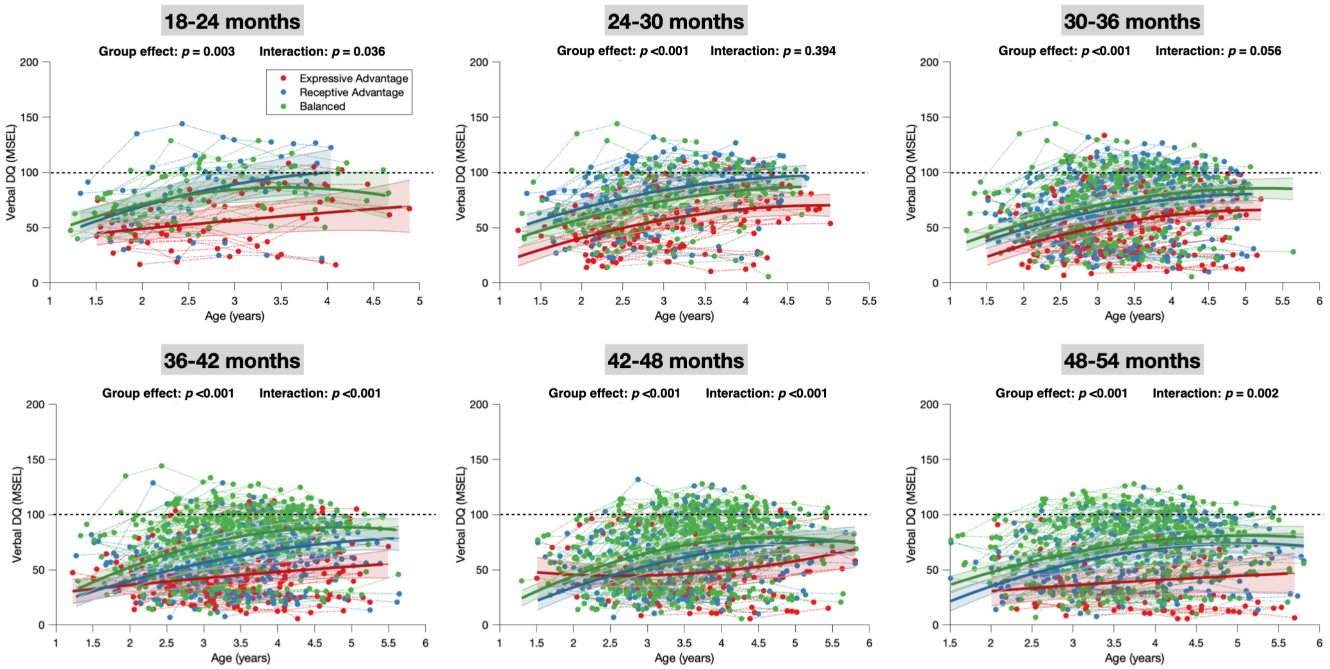


**Figure S6.** Trajectories of verbal development quotient (i.e., average of expressive and receptive language developmental quotients) of the Expressive Advantage, Receptive Advantage, and Balanced profiles in the ASD group across six age windows.

The colored bands around the estimated group-level trajectory indicate the 95% confidence interval. MSEL : Mullen Scales of Early Learning ; DQ: Developmental Quotient.

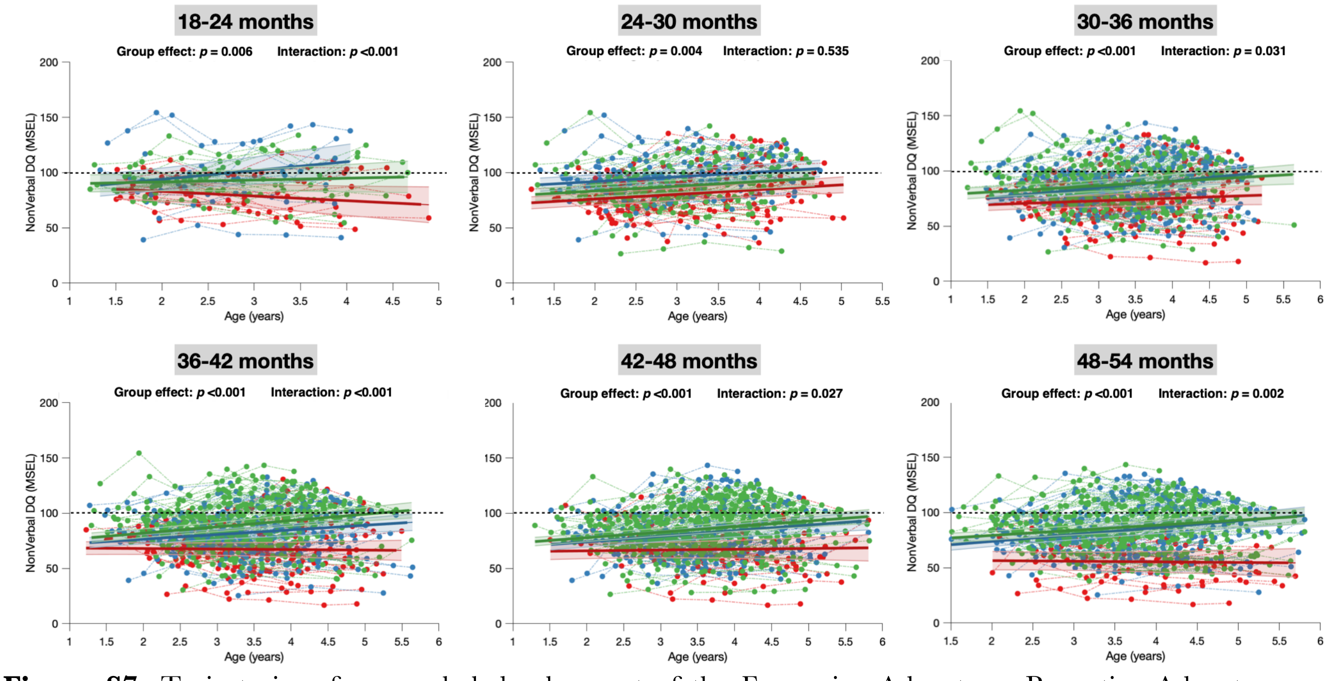


**Figure S7.** Trajectories of non-verbal development of the Expressive Advantage, Receptive Advantage, and Balanced profiles in the ASD group across six age windows.

The colored bands around the estimated group-level trajectory indicate the 95% confidence interval. MSEL : Mullen Scales of Early Learning ; DQ: Developmental Quotient.

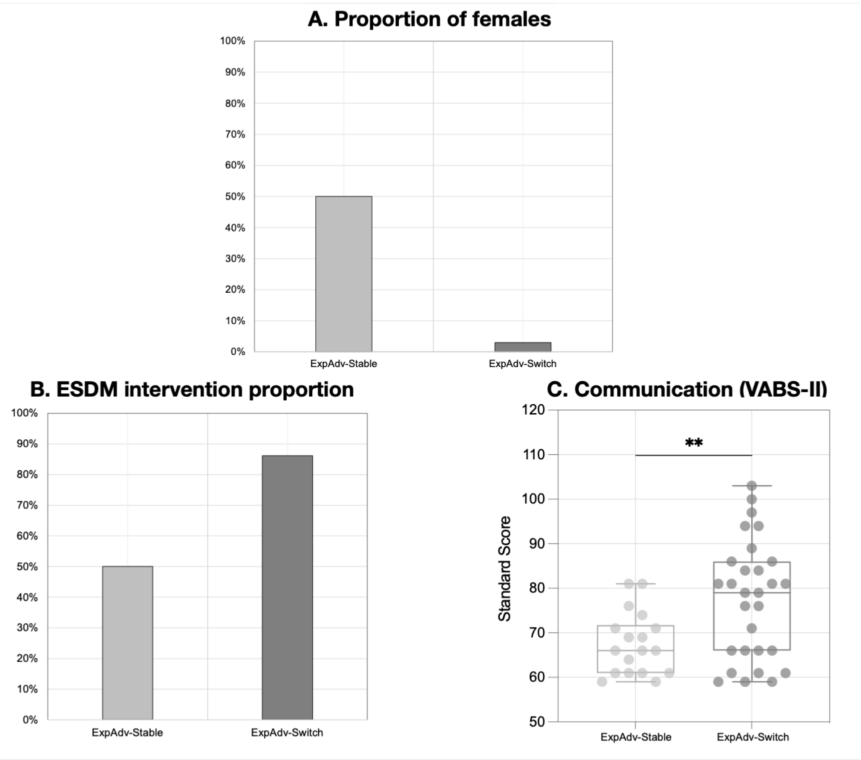


**Figure S8.** Characteristics of autistic children with Expressive Advantage profiles at 30–36 months who either stayed in Expressive Advantage at 36–42 months (n = 18), or who switched into Balanced or Receptive Advantage profiles (n = 29). Panels show: A. Sex Distribution, B. ESDM intervention proportion, C. Communication Standard Score). Sex distribution differed significantly between children who stayed *vs.* those who switched profiles ($p$ <0.001). ESDM intervention proportion also differed ($p$ = 0.017) and children who switched profiles exhibited higher adaptive communication scores than those who stayed in Expressive Advantage (p = 0.006).

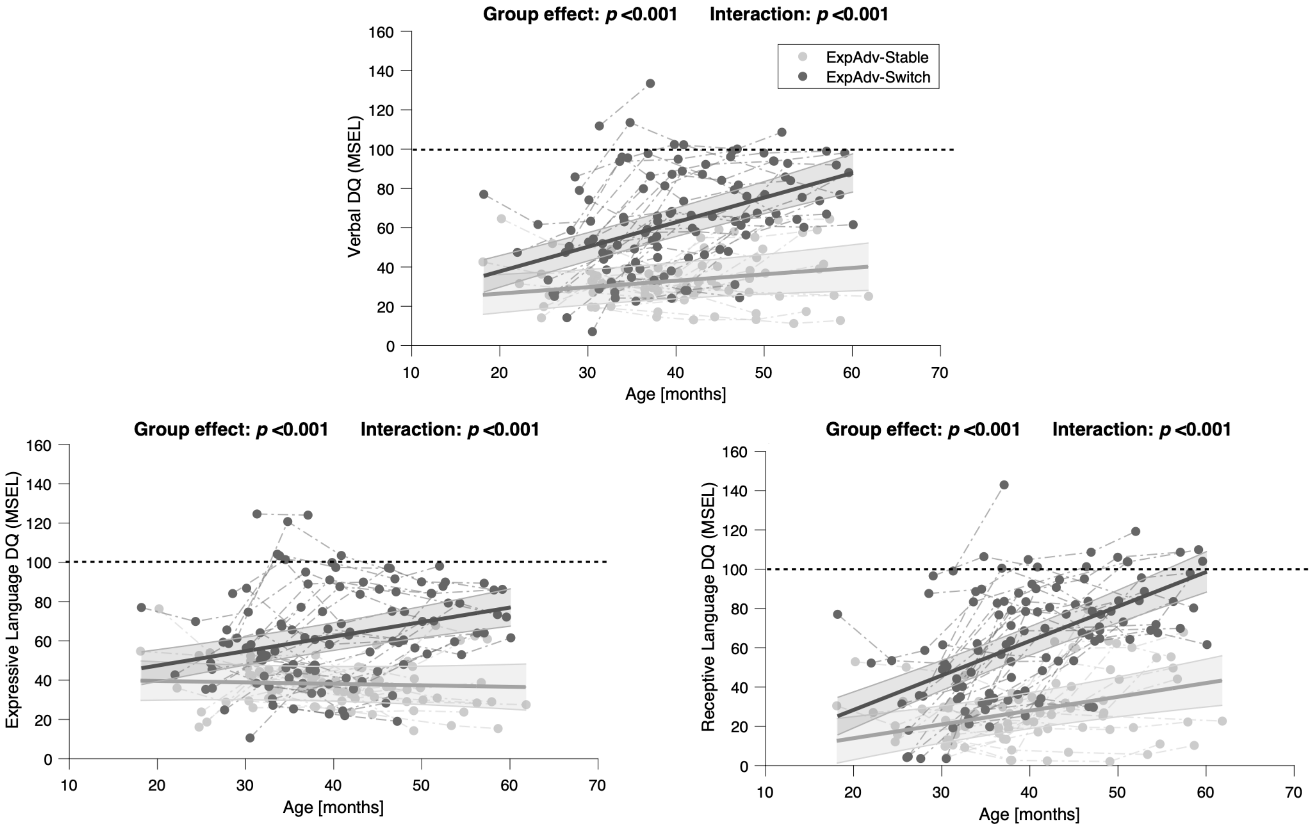


**Figure S9.** Trajectories of Verbal skills, Expressive Language and Receptive Language children classified with Expressive Advantage at the 30–36 month age window, who either then switched profiles into Receptive Advantage or Balanced (yellow, n = 29), or who remained in Expressive Advantage (red, n = 18). profiles in the ASD group across six age windows.

The colored bands around the estimated group-level trajectory indicate the 95% confidence interval. MSEL : Mullen Scales of Early Learning ; DQ: Developmental Quotient. ExpAdv = Expressive Advantage

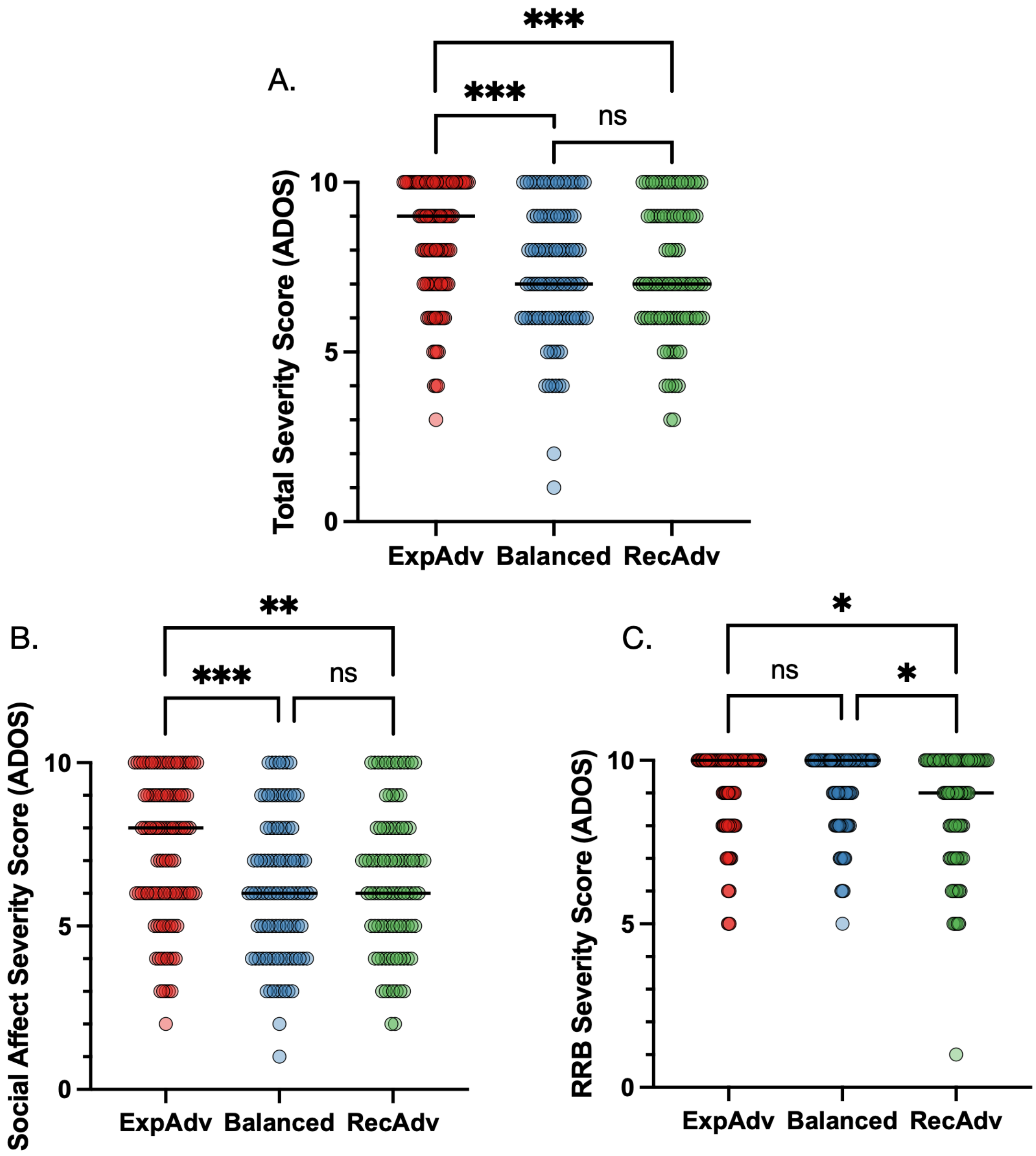


**Figure S10**. Mean ADOS-G/ADOS-2 calibrated severity scores for autistic children in the ExpAdv (red), Balanced (green), and RecAdv (blue) profiles, defined based on the first available time point for each child. (A) Total Calibrated Severity Score; (B) Social Affect Calibrated Severity Score; (C) Restricted and Repetitive Behaviors Calibrated Severity Score.

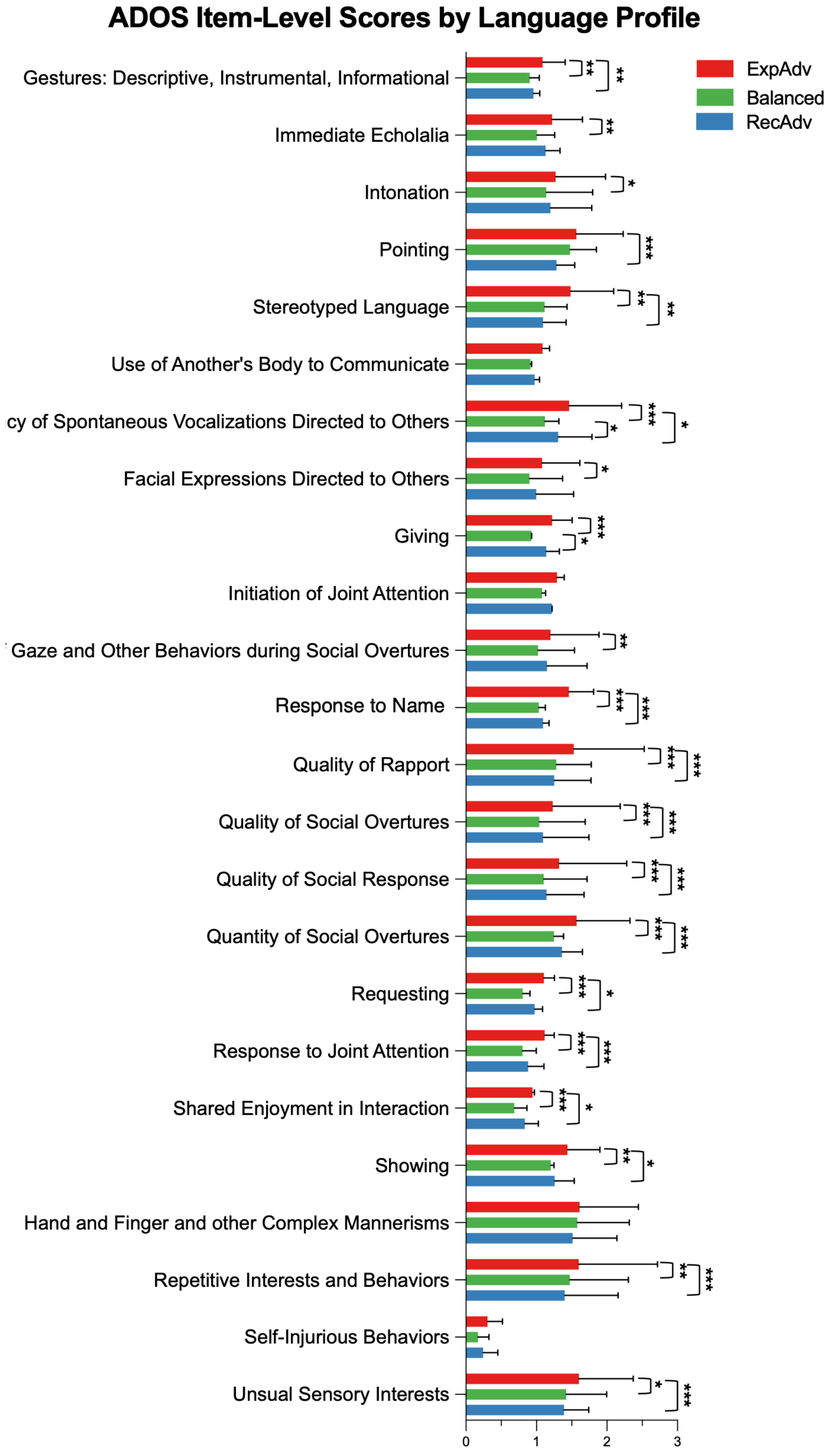


**Figure S11.** ADOS-G/ADOS-2 item-level scores for autistic children in the ExpAdv (red), Balanced (green), and RecAdv (blue) profiles, classified based on the first available time point for each child. Bars represent group median for each item, with horizontal error bars indicating the 95% confidence interval. Item scores range from 0 (no evidence of abnormality) to 3 (clear evidence of abnormality).
